# Integrated Biofluid Proteomics Identified Dynamic Functional Biomarkers of *LRRK2*-Linked Parkinson’s Disease Progression

**DOI:** 10.64898/2025.12.22.25342856

**Authors:** Cong Xiao, Takahiro Shimizu, Bik Tzu Huang, Duc Tung Vu, Ericka Itang, Matthias Mann, Ozge Karayel, Zhenyu Yue

## Abstract

Leucine-rich repeat kinase 2 (LRRK2) variants are the most common cause of inherited Parkinson’s disease (PD), and the hyperactivity of the LRRK2 variants represent a validated drug target for PD. The penetration of common LRRK2 variants is incomplete, underscoring the need for molecular biomarkers that predict disease onset and guide therapeutics development. Here, we analyzed large datasets of cerebrospinal fluid (CSF) and urinary proteomics from the Parkinson’s Progression Markers Initiative (PPMI) and identified distinct lysosomal and immune protein signatures as potential biomarkers for LRRK2-linked PD (LRRK2 PD). Longitudinal analysis revealed that levels of specific lysosomal and immune proteins remained elevated in CSF during the prodromal phase but declined following clinical symptom onset. Furthermore, examination of multiple brain cell types from *Lrrk2* mutant mice carrying disease variant (G2019S) showed heightened secretion of lysosomal proteins in microglia and astrocytes, but not neurons, supporting a glial origin and intrinsic LRRK2 mutant activity responsible for the elevated CSF lysosomal proteins. Furthermore, proteomics analysis of urine from humanized *LRRK2^G2019S^*transgenic mice identified lysosome and glycosphingolipid protein signatures shared with human LRRK2 PD patients. Collectively, our integrated proteomics reveals dynamic changes of functional biofluid signatures for LRRK2 PD, which enables the determination of biomarkers for early disease onset. The humanized *LRRK2*^G2019S^ mice provide a valuable platform for biomarker refinement and therapeutic development.

**One Sentence Summary:** Integrated human and mouse proteomic analyses identify dynamic lysosomal and immune biofluid signatures, possibly of glial origin, as functional biomarkers of LRRK2-linked Parkinson’s disease progression, supported by a novel humanized *LRRK2*^G2019S^ mouse model that recapitulates key urinary biomarker profiles.

## INTRODUCTION

Parkinson’s disease (PD) is the second most common neurodegenerative disorder and characterized by the accumulation of α-synuclein in the form of Lewy bodies and the progressive loss of dopaminergic neurons in the substantia nigra^1,2^. Although advances in diagnostic modalities have improved the clinical detection of PD, a significant number of cases are still under- or mis-diagnosed, especially in early-stage PD^3^. Therefore, there is an urgent need for biomarkers that not only enable early diagnosis of PD but also monitor progression to facilitate therapeutic development.

A major challenge of developing reliable biomarkers for PD is the unclear molecular mechanism and heterogeneity of the disease conditions. Mutations in leucine-rich repeat kinase 2 (*LRRK2*) are the most common cause of autosomal dominant PD^1,4,5^ and have also been linked to sporadic PD (sPD)^6^.

Clinically, LRRK2-associated PD (LRRK2-PD) closely resembles sPD^1^, although Lewy pathology is absent in a subset of cases^7^. Consistent with this neuropathological heterogeneity, a considerable proportion of LRRK2 PD patients were tested negative in α-synuclein seed amplification assays (SAA), which detects α-synuclein oligomers or fibrils with seeding activity and has increasingly been utilized for diagnostic purposes^8,9^. The evidence suggests that in LRRK2 PD, where α-synuclein pathology may be absent or atypical, SAA-based approaches may not be universally applicable and alternative biomarkers are needed to predict the disease onset and progression^8,9^.

Several studies reported specific lysosomal proteins and phospholipids as potential biomarkers for LRRK2 PD. For instance, elevated lysosomal proteins, such as cathepsins, detected in the cerebrospinal fluid (CSF) and urine of individuals carrying *LRRK2* mutations^10–12^. In parallel, levels of bis (monoacylglycerol) phosphate (BMP), a lipid enriched in endolysosomal membranes, are also increased in urine of *LRRK2* mutation carriers^13,14^. Moreover, a recent study identified elevated levels of cytokines and chemokines, such as CCL3, CCL4, CXCL10, and IL-8, in the CSF and plasma of both manifesting and non-manifesting *LRRK2* mutation carriers^15^. These findings may reflect known functions of LRRK2 in immune response and maintenance of lysosome homeostasis^16,17^. These biomarkers may help stratify mutation carriers, and urinary lysosomal proteins in particular may serve as pharmacodynamic markers for evaluating LRRK2-targeted therapies^12^. While these investigations have advanced our biomarker development, biomarkers that capture disease onset and progression are poorly characterized. Given the incomplete penetrance of LRRK2 variants, identification of biomarkers for disease onset or symptom transition becomes crucial to the therapeutics targeting LRRK2 PD.

To identify biomarkers related to disease progression in LRRK2 PD, we analyzed the CSF and urinary proteomes from the Parkinson’s Progression Markers Initiative (PPMI) cohorts in depth and found a shared lysosomal signature in the biofluids of *LRRK2* mutation carriers. Remarkably, longitudinal analysis revealed a dynamic change of lysosomal and immune proteins in male patients. Investigations of multiple brain cell types from *Lrrk2* mutant mice support a glial origin and intrinsic LRRK2 mutant activity responsible for the elevated CSF lysosomal proteins in the carriers. Finally, proteomics profiling of urine samples from humanized LRRK2 mutant transgenic lines identified similar lysosome and glycosphingolipid protein signatures shared with human LRRK2 PD. Collectively, our integrated proteomics reveals dynamic changes of functional biofluid biomarkers for LRRK2 PD, which may facilitate the early prediction of disease onset. Moreover, the novel mouse line facilitates the modeling of urinary endolysosomal biomarkers observed in human *LRRK2* variant carriers, offering a valuable platform for robust biomarker identification and therapeutic development.

## RESULTS

### Overview of PPMI PD cohort datasets and transgenic mouse line datasets

Our study includes three human datasets. The first dataset is human CSF proteomic data, including healthy control participants (HC) (male/female 116/68, mean age 60.66 ± 11.52), PD participants (male/female 361/228, mean age 62.14 ± 9.56, LRRK2-/+ 149, GBA1 -/+ 68, sPD 372), Prodromal PD participants (male/female 139/195, mean age 61.54 ± 7.28, LRRK2-/+ 178, GBA1 -/+ 156). The second dataset is human urine proteomic data, including HC participants (male/female 74/66, mean age 61.05 ± 11.28), PD participants (male/female 290/265, mean age 62.85 ± 9.37, LRRK2-/+ 229, GBA1 -/+ 68, sPD 246), Prodromal PD participants (male/female 189/274, mean age 62.30 ± 7.22, LRRK2-/+ 296, GBA1 -/+ 173). The third dataset is human longitudinal CSF proteomic data, including HC participants (male/female 80/45 mean age 61.48 ± 10.88), PD participants (male/female 183/93, mean age 61.26 ± 9.68, LRRK2-/+ 26, GBA1 -/+ 11, sPD 230, PRKN 9), Prodromal PD participants (male/female 41/32, mean age 63.35 ± 6.77, LRRK2-/+ 38, GBA1 -/+ 11, sPD 24) (Table 1). All these data were generated by PPMI and available to the scientific community.

**Table 1.**
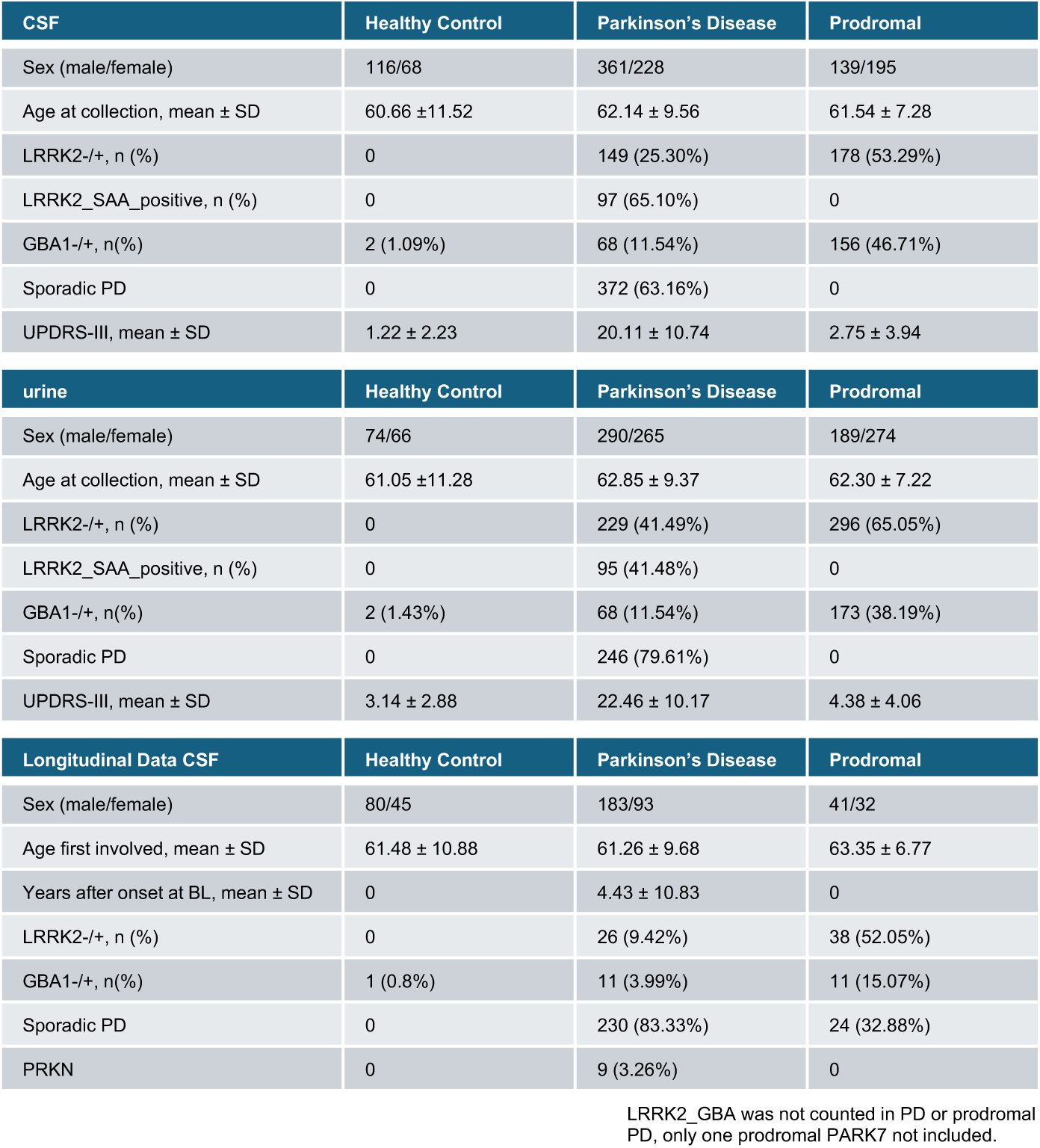
The demographics of all participants.

Additionally, this study includes novel urinary proteomic datasets from transgenic mouse lines, including C57 (WT), *Lrrk2* knock-out (*Lrrk2*^KO^), *Lrrk2*^G2019S^ knock-in (*Lrrk2*^G2019S^), human BAC *LRRK2*^G2019S^ transgenic (hBAC-*LRRK2*^G2019S^), and hBAC-*LRRK2*^G2019S^; *Lrrk2*^KO^ (h*LRRK2*^G2019S^). The overview of our study procedure is presented in Fig. 1a.

**Figure 1.**
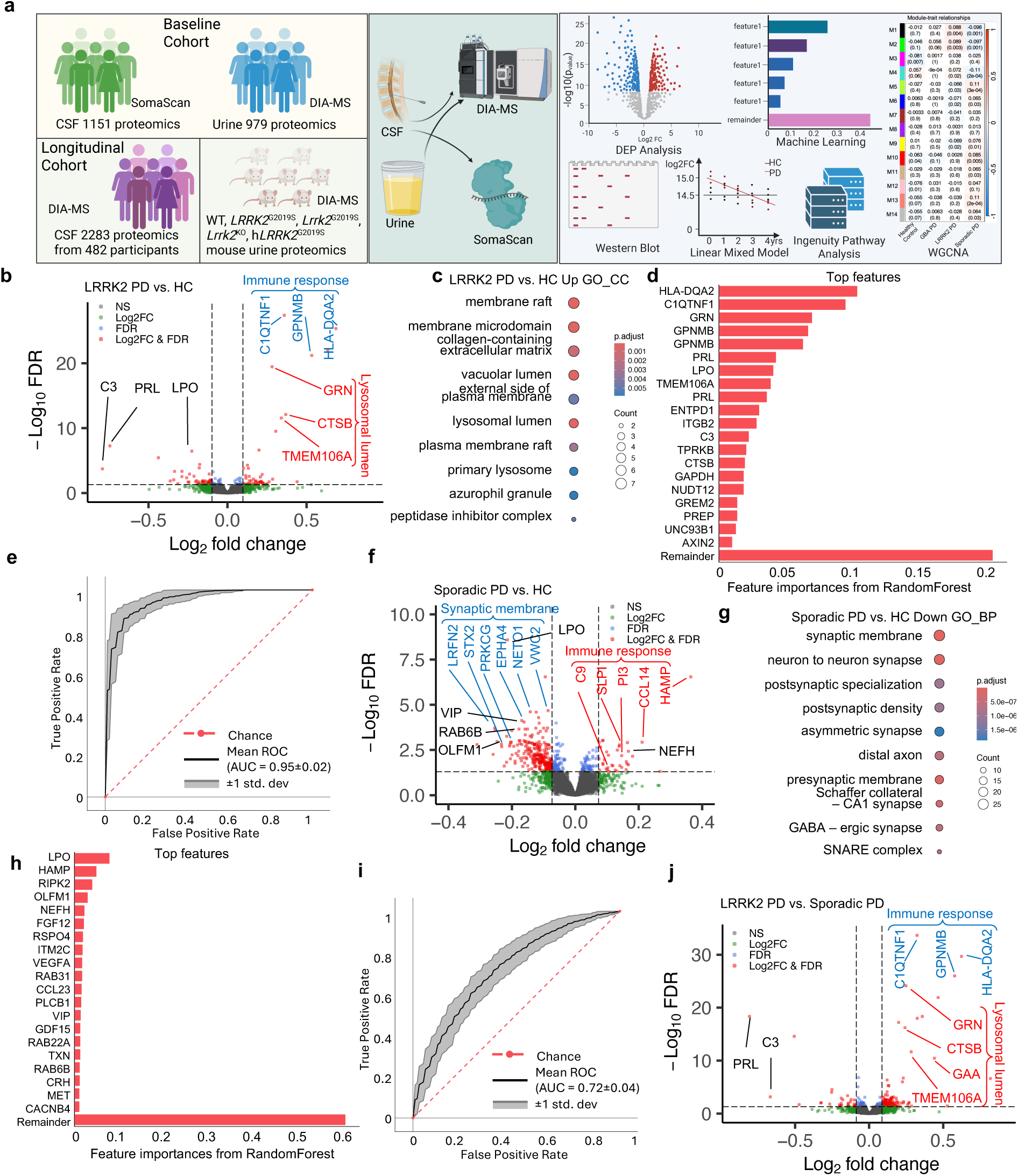
Analysis of CSF proteomics from LRRK2 PD, sporadic PD, and healthy controls. **a,** An overview of the study workflow, which integrates proteomics, bioinformatics, machine learning, cell culture, and protein biochemistry, and investigates multiple baseline CSF and urine, longitudinal CSF and mouse urine proteomic datasets. Created with BioRender.com. **b,f,j** Volcano plot of the differentially abundant proteins (DAPs) detected by mass-spectrometry in human CSF for LRRK2 PD vs. HC (**b**), sPD vs. HC (**d**), and LRRK2 PD vs. sPD (**j**). A positive score indicates enrichment, a negative score indicates depletion. The y axis represents statistical confidence FDR (adjusted P-value) for each x axis point. Enriched proteins, defined by FDR < 0.05 and |Log2FC| > 1.5SD are colored in red. FDR was calculated with the moderated two-sided t-test in the LIMMA package (R Studio) for multiple comparisons. **c,g,** Gene Ontology (GO) annotations of the DAPs in **b (c)** and **f (g)**, displaying the top 10 enriched pathways (*P*-value < 0.05, one-sided hypergeometric test). **d,h**, Feature importance of the top 20 most important features used to distinguish LRRK2 PD vs. HC (d) and (h) individuals in CSF. **e,i**, ROC curve and corresponding AUC statistics in 5-fold cross-validation repeated 10 times using the RandomForest-based model to classify LRRK2 PD vs. HC (**e**) and sPD vs. HC (**i**) based on protein panel in (**d**). Random performance is indicated by the dotted diagonal red line for comparison. Gray area represents the SD from the mean ROC curve. Black lines show the values for a total of repeats with five stratified train-test splits. Red lines show the random chance, corresponding to a classifier that has no discriminative power.

### Identification of lysosomal and immune protein signatures unique to LRRK2 PD through CSF proteomics analysis

We started to explore distinct protein signatures of LRRK2-associated PD (LRRK2 PD) by conducting differentially abundant proteins (DAPs) analysis of the CSF proteomic data across multiple genetically defined PD subgroups (Supplementary Table 1). We found 96 DAPs in LRRK2 PD vs. HC (49 up-regulated, 47 down-regulated, Log2FC > 1.5SD, FDR < 0.05, unless otherwise specified, all analysis were filtered using the same criteria) (Supplementary Table 2), and GO analysis indicated increased pathways associated with lysosome lumen, immune response, and membrane raft, as well as reduced functions in proteasome core complex and endopeptidase complex. The top DAPs in the lysosomal pathway were GRN, CTSB, and TMEM106A; the leading DAPs of immune pathway were C1QTNF1, HLA-DQA2, and GPNMB. The top ranked DAPs of the reduction in LRRK2 PD group were PRL, LPO, and C3 (Fig. 1b,c, and Supplementary Fig. 1a,b). Notably, PRL production can be suppressed by dopaminergic stimulation^18^ and LPO were reported elevated in sporadic PD but decreased in association with higher levodopa equivalent daily doses (LEDD)^19^. We noticed that the LRRK2 PD cohort used in our study had relatively high LEDD, raising the possibility that these differences may reflect treatment effects rather than disease-specific mechanisms^19,20^.

To identify the top features distinguishing LRRK2 PD from HC, we used the open-source machine learning tool OmicLearn^21^. We employed the Extra-Trees package and the 30 most discriminating proteins for training the model within each iteration. These feature proteins were ranked by the classifier based on their feature importance (Fig. 1d). Strikingly, these top features were strongly overlapped with the DAPs enriched in immune (HLA-DQA2, C1QTNF1, and GPNMB) and lysosomal (GRN, TMEM106A, and CTSB) pathways. Additionally, PRL, LPO, and C3 were also among the top features. When these features were employed to train the RandomForest model utilizing the cross-validation technique, we obtained a receiver operating characteristic (ROC) with an average area under the curve (AUC) of 0.95 (standard deviation (std) = 0.02) and a precision-recall (PR) Curve with an average AUC of 0.94 (std = 0.03) (Fig. 1e, and Supplementary Fig. 1c).

In the comparison of sPD vs. HC, we found 301 DAPs (35 up-regulated, 266 down-regulated) (Supplementary Table 3), in which immune response pathway was enriched, while synapse related pathways were reduced (Fig. 1f,g, and Supplementary Fig. 1d). The top feature proteins identified by OmicLearn also showed a strong overlap with DAPs, achieving a ROC with an average AUC of 0.72 (std = 0.04) and a PR Curve with an average AUC of 0.82 (std = 0.03) (Fig. 1h,i, and Supplementary Fig. 1e).

Comparing LRRK2 PD vs. sPD, we found 168 DAPs (136 up-regulated, 32 down-regulated) (Supplementary Table 4), which were enriched in lysosomal (GRN, CTSB, TMEM106A, and GAA) and immune (C1QTNF1, GPNMB, and HLA-DQA2) pathways (Fig. 1j, Supplementary Fig. 1f,g). We next extended our investigation to the proteomics data in GBA PD and found 183 DAPs in GBA PD vs. HC (139 up-regulated, 44 down-regulated) (Supplementary Table 5) and 310 DAPs in GBA vs. sPD (280 up-regulated, 30 down-regulated) (Supplementary Table 6). In both the above comparisons, we observed enriched pathways of immune response and RNA localization, whereas PRL and WIF1 were the top proteins of reduction (Supplementary Fig. 1h-k). This analysis identified lysosomal and immune protein signatures unique to LRRK2 PD.

### Analysis of CSF proteomics across disease phases and SAA+/- subgroups of *LRRK2* and *GBA* variant carriers

To identify potential biomarkers associated with PD onset, we analyzed DAPs between the prodromal and clinical PD phases in both the LRRK2 and GBA cohorts. Only 11 DAPs were detected in comparing LRRK2 PD vs. ProLRRK2 PD (0 up-regulated, 11 down-regulated) (Supplementary Table 7), with PRL, LPO, and VIP as top ranked (Supplementary Fig. 2a). Notably, the lysosomal and immune signature proteins associated with LRRK2 PD were not changed. The analysis of GBA PD vs. proGBA PD also identified a small number of DAPs (10 DAPs, 2 upregulated, 8 downregulated) (Supplementary Table 8), with PRL and LPO as overlapping proteins between the two studies (Supplementary Fig. 2b).

Unlike sPD, LRRK2 PD frequently lacks Lewy pathology, and a fraction may instead be accompanied by tau or TDP-43 accumulation^22^. To explore whether the presence of Lewy pathology influences CSF proteomic profiles within LRRK2 PD, we compared the SAA-positive (+) and SAA-negative (-) subgroups, as SAA positivity represents the presence of pathological α-synuclein “seed” and is often considered a proxy for Lewy pathology^23^. Again, we did not detect any significant DAPs comparing SAA- and SAA+ subgroups in the LRRK2 PD (Supplementary Fig. 2c) (Supplementary Table 9).

Together, the comparison between SAA+ and SAA- subgroups within the LRRK2 PD cohort revealed minimal differences in CSF proteomic profiles. Importantly, the lysosomal and immune protein signatures previously found elevated in LRRK2 PD compared to healthy controls (Fig. 1b,j) were not changed in the above comparisons, suggesting that the increase occurs already at the prodromal stage and is likely independent of Lewy pathology.

### Dynamic changes of lysosome and immune proteins in LRRK2 PD

We next analyzed longitudinal CSF datasets of LRRK2 PD through linear mixed effect model. Initial analysis revealed pronounced differences in the longitudinal trajectories of lysosomal and immune proteins in CSF between sexes (Supplementary Table 10). After partitioning the data by sex, we observed that in males, lysosomal and immune proteins, including CTSD, LAMP2, GM2A, NPC2, PSAP, CTSB, CLU, CD14, B2M, ICOSLG, SERPING1, and CSF1R, remained relatively stable over time in both the LRRK2 prodromal and HC groups, with consistently higher levels in the LRRK2 prodromal group. However, in LRRK2 PD male group these proteins showed a gradual decline across longitudinal timepoints, with some proteins eventually falling below the levels observed in the HC group at later stages. Similarly, extracellular matrix proteins such as EFEMP1, APOE, FN1, CHI3L, TGFBI, and FBLN5 showed comparable levels between the prodromal and HC groups but declined over time in the LRRK2 PD group (Fig. 2a–c and Supplementary Fig. 3a) (Supplementary Table 11).

**Figure. 2.**
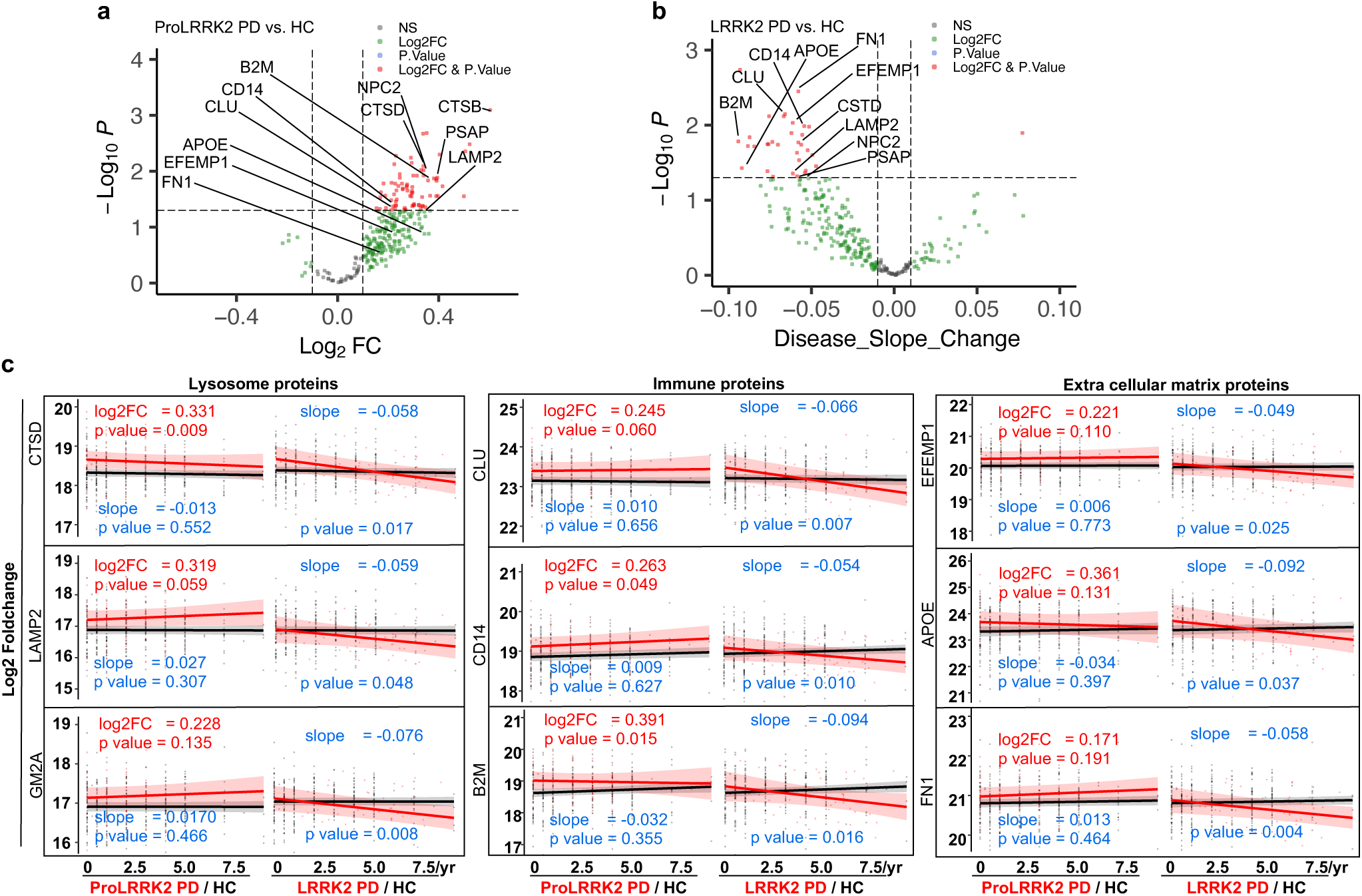
Analysis of longitudinal CSF datasets of LRRK2 PD and healthy control in males. **a**, Volcano plot of the significant enriched proteins at baseline detected by mass-spectrometry in human CSF for prodromal LRRK2 PD vs. HC in males. A positive score indicates enrichment, a negative score indicates depletion. The y axis represents statistical confidence *P*-value for each x axis point. Enriched proteins, defined by *P*-value < 0.05 and |Log2FC| > 0.1 are colored in red. *P*-value was calculated with the moderated two-sided t-test in the LIMMA package. **b**, Volcano plot of the proteins with significant slopes detected by mass-spectrometry in human CSF for LRRK2 PD vs. HC in males. A positive score indicates an increase, a negative score indicates a decrease. The y axis represents statistical confidence *P*-value for each x axis point. Proteins with significant slope change, defined by *P*-value < 0.05 and |slope| > 0.01 are colored in red. *P*-value was calculated with the moderated two-sided *t*-test in the LIMMA package. **c**, Dot plots of longitudinal trajectories of selected protein abundances across disease progression in males. Each dot represents the specific protein expression in an individual sample. The red line denotes the estimated fixed-effect trajectory for prodromal LRRK2 PD or LRRK2 PD, and the black line represents HC. Shaded ribbons indicate the 95% confidence intervals (CIs) of the fixed-effect mean trajectories. Corresponding log2FC, slopes, and p-values are shown alongside each trajectory.

In females by contrast, the lysosomal and immune proteins showed little change through the different disease phases, which might due the limited sample size and proteomic coverage (Supplementary Table 12). Instead, alterations were detected in lipid transport (APOC1, APOC2, APOC3) and acute phase proteins (SERPINA3, SERPINA6, and SERPING1) during the prodromal or PD phases (Supplementary Fig. 3b-d) (Supplementary Table 13).

Together, our analysis revealed the temporal dynamics of protein changes, particularly with lysosomal and immune proteins showing a progressive decline in the PD group, contrasting with the stable profiles observed in the HC and LRRK2 prodromal groups.

### Enhanced sensitivity of lysosomal secretion in *Lrrk2* mutant glia

We hypothesize that the increased levels of lysosomal proteins observed in the CSF of LRRK2 PD patients reflected elevated release of lysosomal proteins from brain cells, given the reported role of LRRK2 in maintaining lysosomal homeostasis^24^. To test the hypothesis, we examined lysosomal protein secretion in primary cells derived from wild-type (WT) and *Lrrk2*^G2019S^ knock-in mice. *Lrrk2*^G2019S^ astrocytes in culture and found a remarkable increase of mature Cathepsin B in medium upon treatment of chloroquine (CQ), compared to WT controls (Fig. 3a,d). A similar increase was also observed for Cathepsin D in the medium of *Lrrk2*^G2019S^ astrocytes (Supplementary Fig. 4a,b). A comparable elevation of Cathepsin B secretion was observed in primary microglia derived from *Lrrk2*^G2019S^ mice following CQ treatment (Fig. 3b,e). In contrast, this enhancement of lysosomal secretion was not observed in primary neurons from *Lrrk2*^G2019S^ mice under the same experimental conditions (Fig. 3c,f). LDH activity in the culture medium did not differ between groups, confirming lack of cytotoxicity (Supplementary Fig. 4c-e). Furthermore, proteinase K treatment reduced the amount of Cathepsin B in the medium, and the combined treatment with detergent and proteinase K did not result in further reduction (Supplementary Fig. 4f,g), suggesting that the secreted lysosomal enzyme is present predominantly in a unencapsulated form. Thus, the above evidence supports a notion that the elevated lysosomal proteins observed in the CSF of LRRK2 PD patients are likely due to the intrinsic LRRK2 mutant function that promotes glial lysosomal secretion.

**Figure 3.**
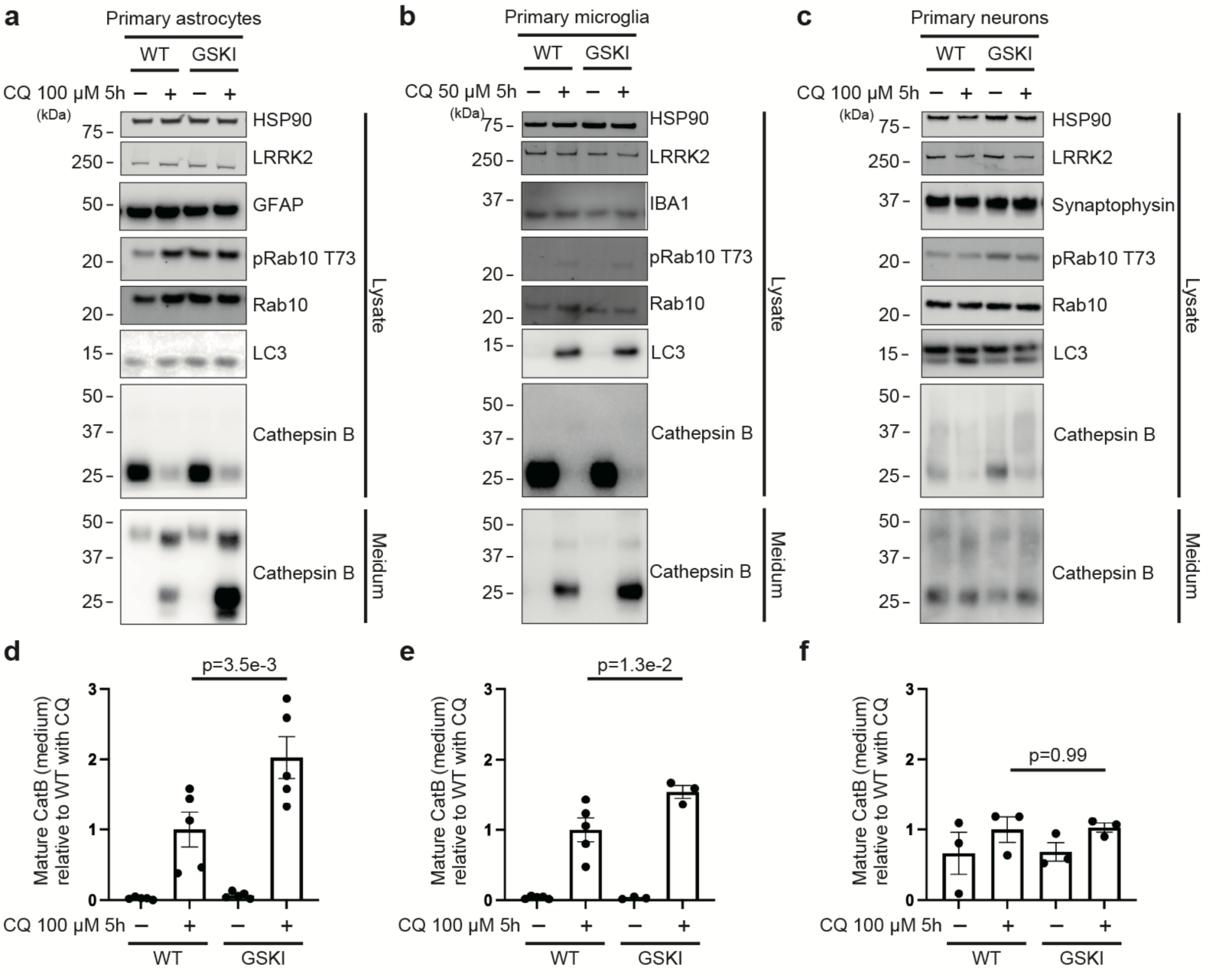
Immunoblot analysis of lysosomal protein secretion from Lrrk2 mutant glia and neurons. **a,b,c,** Immunoblot analysis of lysates and conditioned media from primary astrocytes (**a**), primary microglia (**b**), or primary neurons (**c**) derived from wild-type (WT) or *Lrrk2*^G2019S^ knock-in (GSKI) mice, treated with or without chloroquine (CQ) at the indicated concentrations for 5 hours. Astrocytes were prepared from five independent WT or GSKI pups (n = 5 per group), microglia from five WT and three GSKI pups (WT n = 5, GSKI n = 3), and neurons from three WT and three GSKI pups (n = 3 per group). Immunoblotting was performed using the indicated antibodies. **d,e,f,** Quantification of mature Cathepsin B (observed at around 25 kDa) levels in the conditioned media from astrocytes (**d**), microglia (**e**), and neurons (**f**), corresponding to the immunoblots shown in (**a-c**), respectively. Cathepsin B levels were normalized to the average value of the WT +CQ condition in each cell type. Each data point represents an independent biological replicate derived from a different animal. Bars represent mean ± SEM. Statistical significance was determined by one-way ANOVA followed by Holm–Sidak post hoc test.

### Lysosome and glycolipid metabolism protein signatures revealed from urinary proteomics analysis in LRRK2 PD

Several studies demonstrated that urine showed considerable promise as an alternative biofluid in PD (particularly for LRRK2 PD) for early detection and biomarker discovery^10–12,25,26^. We next analyzed urine proteomics data (PPMI) from human LRRK2 PD vs. HC and observed the enrichment of lysosome lumen pathway (TMEM106B, CTSO, CTSZ, and GBA) and glycolipid metabolic process (GLB1, FUCA1, NAGA, SMPD1, and GM2A); in contrast, the immune response pathway (WFDC2, CD59, CD83, DEFB1, and FCER2) was decreased (Fig. 4a,b and Supplementary Fig. 5a,b) (Supplementary Table 14). The identified feature proteins by OmicLeran showed a strong overlap with the DAPs, achieving a ROC with an average AUC of 0.92 (std = 0.03) and a PR Curve with an average AUC of 0.94 (std = 0.03) (Fig. 4c,d, and Supplementary Fig. 5c). Comparing the urine proteomics of LRRK2 PD vs. sPD, we observed similar patterns of elevated lysosome lumen and glycolipid catabolic pathways in LRRK2 PD (Fig. 4e,f, and Supplementary Fig. 5d,e) (Supplementary Table 15). The analysis of urine data of LRRK2 PD vs. Pro LRRK2 PD identified only a few DAPs, including HBD, HBB, HBA1, AKR1B1, SFRP1, and SDSL (Supplementary Fig. 5f-g) (Supplementary Table 16). No DAPs were observed in the analysis of LRRK2 PD SAA+ vs. LRRK2 PD SAA- subgroups (Supplementary Fig. 5h) (Supplementary Table 17).

**Figure 4.**
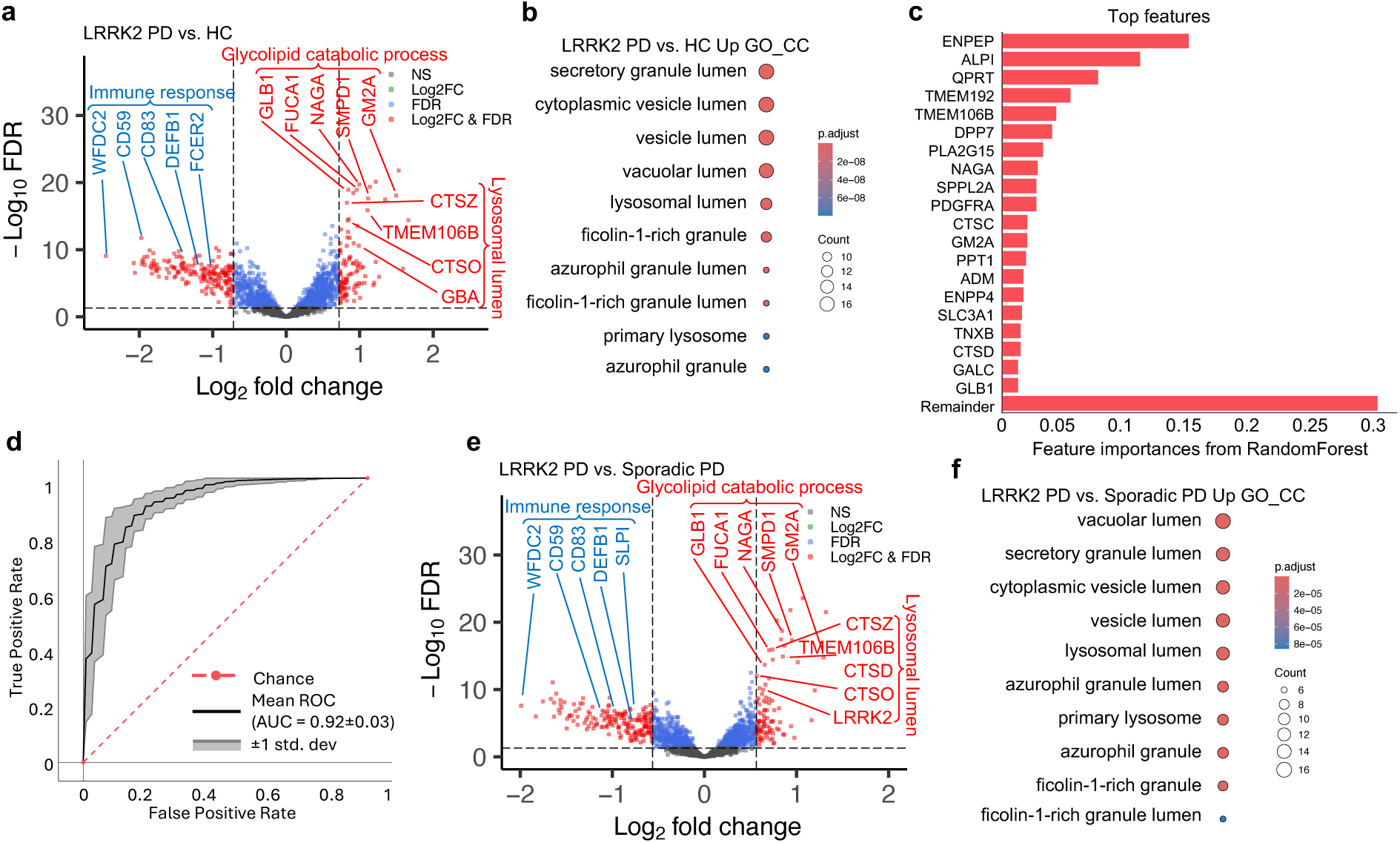
Analysis of urinary proteomics from LRRK2 PD, sporadic PD, and healthy controls. **a**,**e,** Volcano plot of the differentially abundant proteins (DAPs) detected by mass-spectrometry in human urine for LRRK2 PD vs. HC (**a**) and LRRK2 PD vs. sPD (**e**). A positive score indicates enrichment, a negative score indicates depletion. The y axis represents statistical confidence FDR (adjusted *P*-value) for each x axis point. Enriched proteins, defined by FDR < 0.05 and |Log2FC| > 1.5SD are colored in red. FDR was calculated with the moderated two-sided *t*-test in the LIMMA package for multiple comparisons. **b,f,** Gene Ontology (GO) annotations of the DAPs in **a (b)** and **e (f)**, displaying the top 10 enriched pathways (*P*-value < 0.05, one-sided hypergeometric test). **c**, Feature importance of the top 20 most important features used to distinguish LRRK2 PD vs. HC individuals in urine. **d**, ROC curve and corresponding AUC statistics in 5-fold cross-validation repeated 10 times using the RandomForest-based model to classify LRRK2 PD vs. HC based on protein panel in (**c**). Random performance is indicated by the dotted diagonal red line for comparison. Gray area represents the SD from the mean ROC curve. Black lines show the values for a total of repeats with five stratified train-test splits. Red lines show the random chance, corresponding to a classifier that has no discriminative power.

We also analyzed the urine proteomics from GBA and sPD cohorts and investigated DAPs from the comparisons of GBA PD vs. HC, GBA PD vs. sPD, and GBA PD vs. ProGBA PD, as well as sPD vs. HC. Surprisingly, the above comparisons revealed no significant DAPs, due to the same stringent threshold used in our CSF data (FDR and 1.5*SD) (Supplementary Fig. 5i-l). Collectively, the urinary proteomics analysis identified lysosomal lumen and glycolipid metabolic signatures unique to LRRK2-PD (Supplementary Table 18-21).

### Overlapping lysosomal protein signatures between CSF and urinary proteomics of LRRK2 PD

The above results suggest shared lysosomal protein signatures between CSF and urine of LRRK2 PD. To validate the notion, we employed the modified Weighted-Gene-Correlation Network Analysis (WGCNA)^29^, which provides a systematic insight of the relationship among protein networks affected by LRRK2 mutant. In the human CSF data, we identified 14 modules, among which modules M1, M2, and M4 were top ranked and positively correlated with the LRRK2 PD with the *p*-values of 0.004, 0.003, and 0.02, respectively. The lysosomal proteins in M4 were highly ranked by Peptide Significance (PS) and Module Membership (MM), including TMEM106A, CTSH, CTSB, CTSO, GNS, and CTSZ (Supplementary Table 22). The GO term analysis revealed that M4 was enriched in extracellular matrix, early endosome, and lysosomal lumen (Fig. 5a-c). The protein-protein interaction (PPI) analysis of the top 100 proteins of M4 (ordered by PS) through STRING^30^ showed that CTSB, CTSH, PSAP, VIP, and ENTPD1 were connected in the network (Fig. 5d), which explains why they were together among the top features from machine learning results (Fig. 1d). Moreover, M1 is enriched in endosome and vesicle pathways and M2 is enriched in GTPase motor activity and NAD binding (Supplementary Fig. 6a,b).

**Figure 5.**
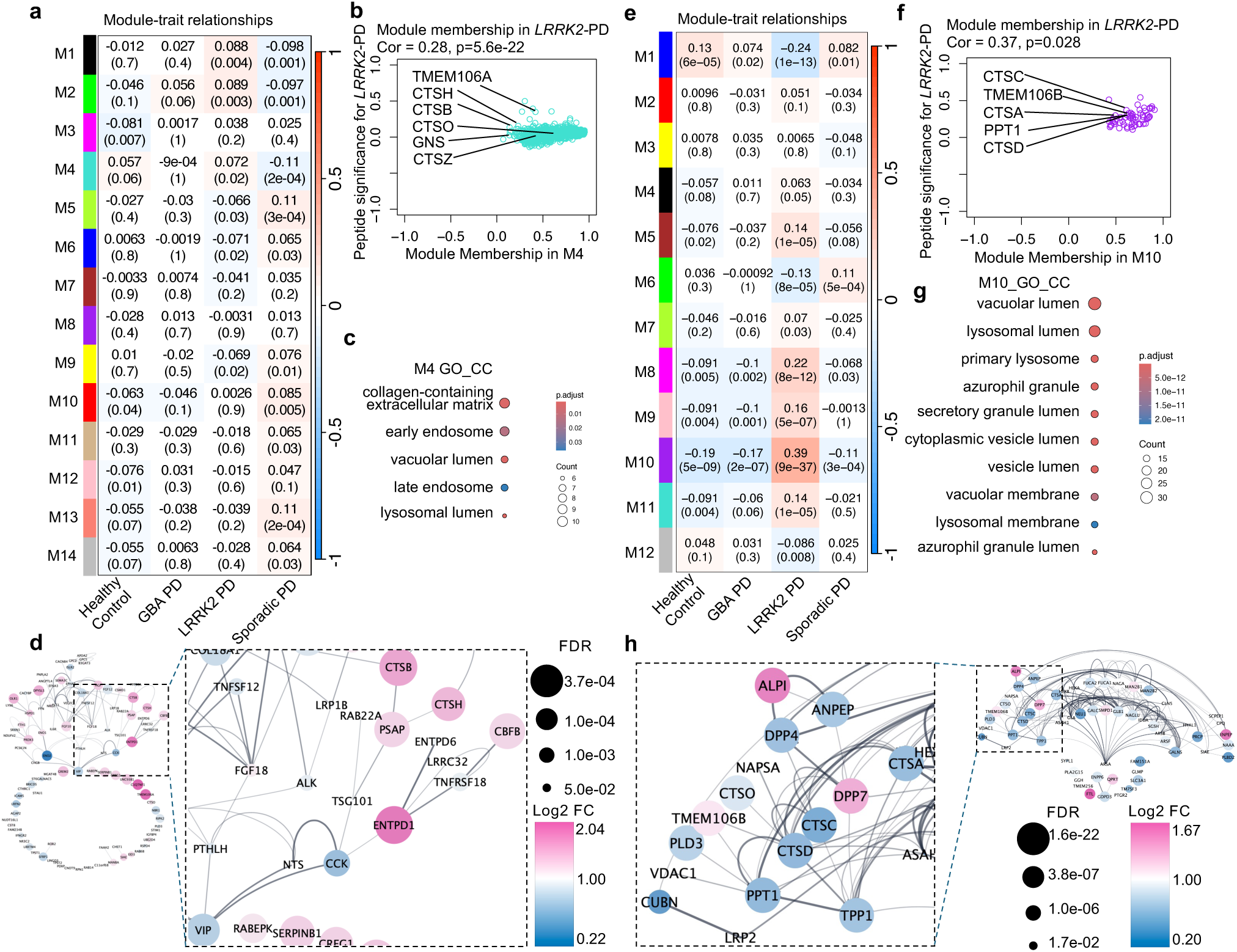
Shared protein signatures between CSF and urinary proteomics of LRRK2 PD. **a,e,** Heatmap of the correlation between the module eigengenes and traits in HC, GBA PD, LRRK2 PD, and sPD in human CSF (**a**) and urine (**d**). Red represents positive correlation, and blue represents negative correlation to the trait. The *Pearson* correlation and two -sided *t*-test *P*-value are presented in each module. **b,f,** Scatter plot of the module membership (x axis) and peptide significance (y axis) in M4 from human CSF with *P*-value < 0.02 (b) or M10 from human urine with *P*-value < 9e-37 (f). *P*-value was calculated with two-sided *t*-test. **c,g,** Gene Ontology (GO) annotations of the proteins in M4 from human CSF (**c**) or M10 from human urine (**f**), displaying the top 10 enriched pathways (*P*-value < 0.05, one-sided hypergeometric test). **d,h,** PPI network analysis of proteins in M4 in a (top 100 proteins ordered by PS). The color gradience represents from low (blue) to high (pink) fold enrichment. Dot size represents statistical significance. FDR was calculated with the moderated two-sided t-test in the LIMMA package for multiple comparisons.

In the human urine data, we identified 12 modules, among which the top three modules M5, M8, and M10 (*p*-values of 1e-05, 8e-12, and 9e-37, respectively) shared similar pathways with the top three modules in CSF (M1, M2 and M4). The lysosomal proteins were highly ranked (significant PS and MM) in M10 modular, including CTSS, TMEM106B, CTSA, CTSB, and TPP1 (Supplementary Table 23).

Lysosome lumen, lysosomal membrane, sphingolipid metabolic process, glycolipid catabolic process, and glycosphingolipid catabolic process were significantly enriched in M10 through GO term analysis (Fig. 5 e-g and Supplementary Fig. 6c). The PPI analysis of the proteins in M10 showed that the interconnectivity of the lysosomal proteins, particularly PPT1, CTSD, CTSC, TPP1, which were in the center position of the network (Fig. 5 h). The majority were also among the top features from machine learning results (Fig. 4c). Similar to the M1 and M2 in CSF, the M5 and M8 from urine dataset were enriched in vesicle lumen and secretory granule lumen, ATP hydrolysis activity and GTP binding, respectively (Supplementary Fig. 6d,e). The WGCNA from the proteomics dataset revealed shared lysosomal protein signatures between CSF and urine of LRRK2 PD proteomics, while glycosphingolipid protein signature is urine specific.

We next assessed the cross-biofluid expression concordance of a curated lysosomal core protein panel^31^ in CSF and urine from the same individuals. Among 137 matched LRRK2 PD participants in both biofluids, 45 lysosomal core proteins were consistently detected. *Pearson* correlation analysis revealed that 28 proteins exhibited positive, whereas 18 showed negative correlations between CSF and urine abundances, even though they were generated by different platforms (SomaScan and MS). Among these, MAN2B2, IDS, and CTSB displayed the highest positive correlation coefficients (Supplementary Fig. 6f,g) (Supplementary Table 24).

### Urinary proteomics analysis of mouse models expressing human *LRRK2*^G2019S^ associated with lysosome and glycosphingolipid metabolism signatures

To validate human LRRK2 variant-associated biofluid signatures, we sought to investigate LRRK2 PD mutant mouse models. Due to the challenge of obtaining sufficient CSF from mice for proteomics analysis, we focused on urine samples. We first investigated *Lrrk2*^G2019S^ KI and *Lrrk2*^KO^ mouse lines and performed proteomics analysis of their urine samples. We observed 196 DAPs in urine samples (80 up-regulated, 116 down-regulated) in *Lrrk2*^G2019S^ mice vs. WT mice comparison (Supplementary Table 25). The GO analysis indicated an increase in mitochondrial matrix and multiple catabolic processes pathways as well as a decrease in translation at pre-/post-synapse (RPL10a, RPL27, RPL26, RPL4, RPL13, RPL8, PRS14, PRS15A, and PRS11) (Supplementary Fig. 7a-c). However, we saw no change in the lysosomal pathway in *Lrrk2*^G2019S^ mice. In contrast, analysis of urine samples from *Lrrk2*^KO^ vs.

WT mice showed 54 DAPs (17 up-regulated, 39 down-regulated, FDR < 0.05) (Supplementary Table 26) with a significant decrease in the lysosomal lumen (TMEM106B, CTSA, CTSH, PLD3, and GUSB), glycosphingolipid (GSLs) and catabolism (GLB1, GALC, and NEU1) pathways (Fig. 6a,b, Supplementary Fig. 7d,e), consistent with a role of mouse Lrrk2 in promoting the secretion of lysosomal proteins^32,33^.

**Figure 6.**
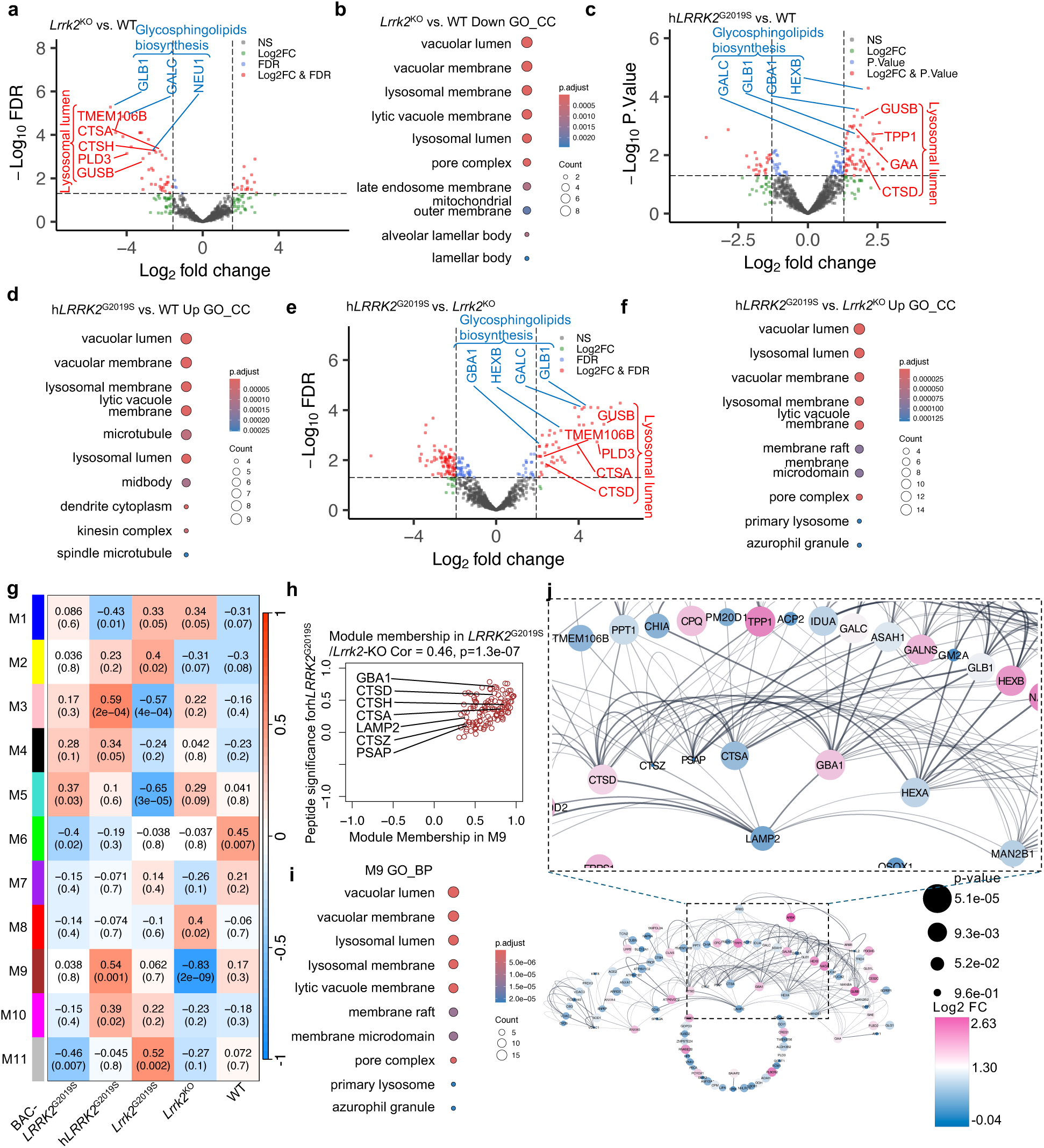
Analysis of urine proteomics from Lrrk2G2019S, Lrrk2KO, BAC-LRRK2G2019S, hLRRK2G2019S, and WT mice. **a,c**,**e,** Volcano plot of the differentially abundant proteins (DAPs) detected by mass-spectrometry in mouse urine for *Lrrk2*^KO^ vs. WT (**a**), h*LRRK2*^G2019S^ vs. WT (**c**), and h*LRRK2*^G2019S^ vs. *Lrrk2*^KO^ (**e**). A positive score indicates enrichment, a negative score indicates depletion. The y axis represents statistical confidence FDR (adjusted *P*-value)/*P*-value for each x axis point. Enriched proteins, defined by FDR/*P*-value < 0.05 and |Log2FC| > 1.5SD are colored in red. FDR/*P*-value was calculated with the moderated two-sided *t*-test in the LIMMA package for multiple comparisons. **b,d,f** Gene Ontology (GO) annotations of the DAPs in **a** (**b**), **c** (**d**), and **e** (**f**) displaying the top 10 enriched pathways (*P*-value < 0.05, one-sided hypergeometric test). **g,** Heatmap of the correlation between the module eigengenes and traits in WT, BAC-h*LRRK2*^G2019S^, h*LRRK2*^G2019S^, *Lrrk2*^G2019S^, and *Lrrk2*^KO^ in mouse urine. Red represents positive correlation, and blue represents negative correlation to the trait. The *Pearson* correlation and two -sided *t*-test *P-value* are presented in each module. **h,** Scatter plot of the module membership (x axis) and peptide significance (y axis) in M9 from mouse urine with *P*-value < 0.001. *P*-value was calculated with two-sided *t*-test. **i,** Gene Ontology (GO) annotations of the proteins in M9 from mouse urine, displaying the top 10 enriched pathways (*P*-value < 0.05, one-sided hypergeometric test). **j,** PPI network analysis of proteins in M4 in a (top 100 proteins ordered by PS). The color gradience represents from low (blue) to high (pink) fold enrichment. Dot size represents statistical significance. p-value was calculated with the moderated two-sided t-test in the LIMMA package.

The lack of observed human urinary lysosome signature in *Lrrk2*^G2019S^ mice could be due to the absence of unique human LRRK2 pathogenic activity. We decided to introduce human genomic LRRK2^G2019S^ sequence into mice due to noticeably differential expression levels of LRRK2 in various cell types and potentially different enzymatic activities between the species^34^. We crossed a human BAC-*LRRK2*^G2019S^ transgenic line to *Lrrk2*^KO^ mice to generate a “humanized” mutant line h*LRRK2*^G2019S^ (Supplementary Fig. 8a). The h*LRRK2*^G2019S^ mouse line shows comparable protein levels of human LRRK2 mutant to WT mouse LRRK2 in tissues however with remarkably elevated phosphorylation of Rab10 and Rab12 (Supplementary Fig. 8b-f). The robust kinase activity of human *LRRK2*^G2019S^ in the h*LRRK2*^G2019S^ tissues contrast with the result from *Lrrk2*^G2019S^ KI line^35^. Proteomic analysis of urine from h*LRRK2*^G2019S^ male mice identified 81 DAPs (58 up-regulated, 23 down-regulated, *p*-value less than 0.05), compared to WT mice (Supplementary Table 27). GO term analysis indicated that the most significantly enriched pathways were lysosomal lumen (GUSB, TPP1, GAA, and CTSD) and glycosphingolipid biosynthesis (GALC, GLB1, GBA1, and HEXB), followed by dendrite cytoplasm and proteoglycan catabolic process (Fig. 6c,d, and Supplementary Fig. 8g,h). Moreover, when analyzing samples from h*LRRK2*^G2019S^ vs. *Lrrk2*^KO^ mice through stringent analysis (FDR < 0.05), we observed 106 DAPs (60 up-regulated, 46 down-regulated) (Supplementary Table 28), of which lysosomal lumen (TMEM106B, CTSA, CTSD, PLD3, and GUSB) and glycosphingolipid biosynthesis (GLB1, CALC, GBA1, and HEXB) pathways were the top ranked enriched pathways, followed by membrane raft, pore complex, vacuole organization, and membrane lipid catabolic process (Fig. 6e,f, Supplementary Fig. 8i,j).

Thus, h*LRRK2* mutant not only was able to functionally rescue the reduction of urinary lysosomal proteins observed in *Lrrk2*^KO^ mice, but also caused enhanced lysosomal protein levels compared to WT. Moreover, the h*LRRK2*^G2019S^ vs. *Lrrk2*^KO^ comparison revealed a robust enhancement of the signature resembling those in human *LRRK2* mutation carriers, suggesting that this model provides a functionally relevant system for studying LRRK2-associated urinary biomarkers.

We next performed WGCNA to identify module-trait relationships by integrating mouse urine data across five different genotypes. The results showed that only M9 module is positively correlated with h*LRRK2*^G2019S^ (*p* = 0.001) and negatively correlated with *Lrrk2*^KO^ (*p* = 2e-09). M9 is significantly enriched in lysosomal proteins (significant PS and MM), including GBA1, CTSD, CTSH, CTSA, LAMP2, CTSZ, and PSAP (Supplementary Table 29). GO term analysis further verified the enrichment in lysosome lumen, lysosome membrane, membrane lipid catabolic process, glycolipid catabolic process, and glycosphingolipid catabolic process pathways, which were similar to the human urine M10 module (Fig. 5e-g, Fig. 6g-i, and Supplementary Fig. 9a). The above network analysis demonstrated that the signature proteins in M9 represent functional biomarkers associated with LRRK2 PD pathogenic pathway in the urine. The PPI analysis of the top 100 proteins from M9 (ordered by PS) showed interconnectivity of the lysosomal proteins, among which HEXA, HEXB, CTSA, CTSZ, CTSD, and LAMP2 were in the center position of this network (Fig. 6 j). Together, our results demonstrated that the urine signatures of h*LRRK2*^G2019S^ mice recapitulate those of human LRRK2 PD samples.

### Functional biofluid signatures of LRRK2 PD from integrated proteomics

We next employed the knowledge-based Ingenuity Pathway Analysis (IPA)^36^ and performed the Comparison Analysis to investigate the signature proteins from human CSF, urine, and mouse urine samples. CLEAR signaling and neutrophil degranulation were among the top-ranked canonical pathways consistently shared across all comparisons analyzed. Proteins associated with these pathways were elevated in the urine of h*LRRK2*^G2019S^ compared to WT or *Lrrk2*^KO^ mice, and in the CSF and urine of LRRK2 PD patients compared to HC, whereas they were decreased in the urine of *Lrrk2*^KO^ mice compared to WT(z-score > 2 or < -2) (Supplementary Fig. 9b) (Supplementary Table 30). In the Upstream Analysis, transcription factor EB (TFEB) was top ranked, showing similar patterns across all comparisons (Supplementary Fig. 9b,c) (Supplementary Table 31). TFEB plays a key role in the regulation of lysosome-associated processes, including autophagy, exo- and endocytosis, phagocytosis and immune response^37^.

## DISCUSSION

By combining large PPMI proteomic datasets from human PD biofluid (CSF and urine) and the urine proteomics from humanized LRRK2-G2019S variant mouse models, our integrated studies identified dynamic functional signatures of *LRRK2*-linked PD progression. These LRRK2 PD signatures are lysosomal and immune-related proteins, which are tied to the intrinsic LRRK2 function and dysregulated by LRRK2 variants. To our knowledge, this study is the first to show elevated lysosomal proteins in the CSF of LRRK2 mutation carriers based on the largest CSF cohort using SomaScan platform, even though several studies reported similar lysosome changes in human CSF within small cohorts of PD patients^10,11,38^.

Our study revealed an increase of the proteins associated with glycolipid/glycosphingolipid (GSL) catabolic process in urine of both LRRK2 PD patients and h*LRRK2*^G2019S^ mice. The observation is consistent with previous report that bis-monoacylglycerol-phosphate (BMP) level was elevated in urine of *LRRK2*^G2019S^ carries ^36,37^, as BMP stimulates lysosomal lipid hydrolases and helps sort/degrade lipid cargo (including GSL)^39^. The increase of immune and lysosome proteins is unique to LRRK2 PD, as evidenced by lack of such signatures in sPD and GBA-PD cohorts. In addition, we observed that *Lrrk2*^G2019S^ sensitized the lysosomal protein secretion from glial cells, which helps explain the enhanced lysosomal markers in CSF. Taken together, the elevated levels of lysosomal proteins in CSF and GSL catabolic process–related proteins in urine from LRRK2 variant carriers may indicate increased turnover and exocytosis of lysosomes driven by hyperactive LRRK2 mutations. Our study thus provides critical insight into the development of functional biomarkers for LRRK2-linked PD.

A key finding of our study is the dynamic changes of lysosomal and immune-related proteins in LRRK2 variant carriers through longitudinal analysis. We found a substantial elevation of lysosomal and immune signature proteins during the prodromal phase, followed by their decline along disease progression, only in males, possibly reflecting the clinicopathological differences of LRRK2 PD between sexes^39^. The prodromal increase of lysosomal and immune signatures in LRRK2 variant biofluid may be tied to LRRK2 intrinsic hyperactivity. However, it remains unclear when the elevation initiates, as the cohort includes only individuals aged 50 or older. Whether such biofluid LRRK2 variant signatures are necessary to drive disease onset and progression in PD is also unclear, as they are absent in female LRRK2 variants, sPD or GBA PD cohorts. Furthermore, the male-specific progressive reduction of the LRRK2 signatures in CSF during disease progression could be related to the recently reported sex- and age-dependent decline of lysosomal functions and immune responses driven by increased LRRK2 kinase activity^17^. Alternative mechanisms underlying the observed decline of lysosomal proteins may involve TFEB, a master regulator of lysosomal biogenesis, as impaired TFEB function could lead to a decrease in intracellular lysosomal content, thereby limiting the amount of protein available for secretion. In fact, Ingenuity Pathway Analysis of the integrated proteomic datasets identified CLEAR signaling as the top shared canonical pathway and TFEB as the top upstream regulator, suggesting dysregulation of this pathway in brain cells. TFEB activity was reported to be reduced in sporadic PD^40^, and its activity is also negatively associated with LRRK2 kinase activity^41^, which may be mechanistically related to the present findings. The relationship between disease progression and TFEB activity in LRRK2 PD, as well as the link between CSF lysosomal protein levels and intracellular TFEB activity, remains to be elucidated and warrants further investigation.

There are some limitations to this study. While the above ideas require to be tested, future studies should address several questions raised by our observations, including the sex- and gene-specific LRRK2 PD signatures, whether the longitudinal decline of LRRK2 signatures precedes or follows the onset of motor symptoms, and the need for longitudinal data from individuals who eventually phenoconvert, as these temporal relationships cannot be directly assessed in our dataset given that the LRRK2 prodromal and PD groups comprise different individuals. A key limitation of our current longitudinal cohort is the relatively small number of LRRK2-PD participants and the restricted proteomic coverage (291 proteins) compared to the CSF baseline dataset. These factors reduce the statistical power of our analyses.

Accordingly, future work will require substantially larger cohorts, broader proteome coverage and deeper sequencing to validate our findings and enhance robustness.

Mechanistically, LRRK2 has been implicated in stress-induced lysosomal exocytosis^24^, although it has remained unclear whether pathogenic mutations enhance this process^42,43^. We found that the G2019S mutation increases lysosomal protein secretion selectively in glial cells, but not in neurons. Notably, the set of lysosomal proteins elevated in LRRK2 mutation carriers included GPNMB, which was recently reported to be highly enriched in glial lysosomes, further supporting our conclusion that these CSF changes likely originate from glial cells^44^. In contrast, recent studies have shown that neurons harboring the LRRK2 G2019S mutation release exosomes enriched in mitochondrial proteins, possibly due to impaired autophagy and aberrant routing of autophagosomal cargo^45^. This mechanism appears distinct from glial lysosomal protein secretion, as our proteinase K protection assay demonstrated that secreted glial lysosomal proteins are not membrane-enclosed, unlike exosomes. Mitochondrial proteins were not enriched in our CSF dataset, potentially due to technical limitations such as their low abundance or inefficient detection by standard proteomic workflows. Nonetheless, as observed for the coordinated trajectory of glia-derived lysosomal and immune-related proteins, neuronal exosome-derived constituents including mitochondrial proteins may also display dynamic longitudinal changes in LRRK2 PD. Elucidating such temporal patterns may help distinguish glial- and neuron-derived biomarkers that capture distinct aspects of disease progression and deepen our understanding of cell-type–specific mechanisms in LRRK2 PD pathogenesis.

Our study reports a novel h*LRRK2^G2019S^* mouse model, which recapitulated the urinary lysosomal protein signature observed in human LRRK2 PD. Notably, this model exhibits robust kinase activity (Supplementary Fig. 8c-f), leading to enhanced phosphorylation of Rab10 and Rab12, consistent with the phospho-Rab profiles reported in the brains of LRRK2 PD with tau pathology^46^. Thus, compared to conventional knock-in mice, the h*LRRK2^G2019S^* model captures key molecular features of the human LRRK2 mutations, providing a valuable platform for investigating biomarker dynamics and their underlying biological mechanisms. Importantly, recent urinary proteomics study in BAC-h*LRRK2*^G2019S^ rats have likewise demonstrated a lysosomal/glycosphingolipid-enriched urinary signature associated with pathogenic LRRK2, and importantly, showed that genetic *Lrrk2* knockout or pharmacological inhibition of LRRK2 kinase activity reverses these alterations in vivo^12^. In an analogous manner, our h*LRRK2^G2019S^* mouse model can serve as an in vivo platform for validating the effects of LRRK2 kinase inhibition. Moreover, because multiple well-characterized pathogenic PD models have been established in mice^47,48^, this system can be readily combined with additional genetic or environmental perturbations to investigate mechanistically how LRRK2-linked urinary biomarkers relate to disease-relevant pathways.

## MATERIALS AND METHODS

### Animals

All animal procedures were conducted in accordance with the guidelines of the Icahn School of Medicine at Mount Sinai Institutional Animal Care and Use Committee (IACUC protocol #: 2021-00031). Mice were housed under a 12-hour light/12-hour dark cycle at a controlled temperature of 20–23 °C and relative humidity of 40–60%, with ad libitum access to food and water. Wild-type C57BL/6J mice (JAX: 000664) were obtained from The Jackson Laboratory. Lrrk2 G2019S knock-in mice (GSKI mice) were kindly provided by Dr. Deanna Benson. Both wild-type and GSKI mice were used for the preparation of primary astrocytes, neurons, and microglia. Humanized LRRK2 G2019S mice (hLRRK2 G2019S) were generated by crossing *Lrrk2* knockout (KO) mice (C57BL/6-Lrrk2<tm1.1Mjff>/J; JAX: 016121) with human BAC transgenic hLRRK2 G2019S mice (C57BL/6J-Tg(LRRK2*G2019S)2AMjff/J; JAX: 018785), both purchased from The Jackson Laboratory. Mice carrying the homozygous Lrrk2 KO and hemizygous human BAC-hLRRK2 G2019S alleles were designated as hLRRK2 G2019S mice.

### Antibodies and reagent

For immunoblotting, the following antibodies were used: anti-HSP90 (61049, BD Transduction Lab), anti-LRRK2 (ab133474, abcam), anti-phosphoLRRK2 (ab203181), anti-GFAP (13-0300, Invitrogen), anti-IBA1 (019-19741, Wako), anti-synaptophysin (101011, Synaptic Systems), anti-phosphoRab10 (ab230161, abcam), anti-Rab10 (ab237703, abcam), anti-phosphoRab12 (ab256487, abcam), anti-Rab12 (sc-515613, Santa Cruz), anti-LC3 (PM036, MBL), anti-Cathepsin B (31718, Cell Signaling), anti-CathepsinD (ab75852, abcam). For fluorescent detection, IRDye® 800CW-conjugated secondary antibodies were used: anti-mouse (926-32210, LI-COR Biosciences), anti-rabbit (926-32211, LI-COR Biosciences), anti-rat (926-32219, LI-COR Biosciences), and anti-guinea pig (926-32411, LI-COR Biosciences). Horseradish peroxidase (HRP)-conjugated anti-mouse immunoglobulin G (111-035-003, Jackson ImmunoResearch Laboratories) and anti-rabbit immunoglobulin G (111-035-144, Jackson ImmunoResearch Laboratories) antibodies were used as secondary antibodies. As described in the figure, 50 or 100 μM Chloroquine (C6628, Sigma Aldrich) was applied.

### Primary microglia/astrocyte culture

Neonatal mice (postnatal day 0–2) were anesthetized by hypothermia on ice, and whole brains were rapidly dissected. The olfactory bulbs, meninges, cerebella, and brain stems were carefully removed, and the remaining cerebral hemispheres were transferred into individual tubes containing ice-cold glial medium. This medium consisted of Dulbecco’s Modified Eagle’s Medium (DMEM; 11965-092, Gibco, Thermo Fisher Scientific) supplemented with penicillin–streptomycin (15070063, Gibco), GlutaMAX (35050061, Gibco), and 20% heat-inactivated fetal bovine serum (FBS; SH30396.03HI, HyClone).

Brain tissue from each pup was processed separately. Tissue dissociation was performed mechanically, first using a 1 mL plastic pipette tip, followed by gentle trituration with a fire-polished glass Pasteur pipette. The resulting single-cell suspensions were plated into T75 flasks and maintained in a humidified incubator at 37 °C with 5% CO₂ for 14 days. On day in vitro 14 (DIV14), adherent astrocytes were detached using 0.05% trypsin–EDTA (25200056, Gibco) and re-plated at appropriate densities onto culture plates for subsequent immunoblotting. Floating microglia were collected from the astrocyte layer by gentle mechanical agitation (tapping the flasks), and the supernatant was passed through a 40 µm cell strainer to remove debris and cell aggregates. Isolated microglia were then plated at appropriate densities for downstream applications, including immunoblot analysis.

### Primary neuron culture

Primary cortical neurons were prepared from embryonic day 17–18 mouse embryos. Following dissection, meninges-free cortices were carefully isolated and enzymatically dissociated in a solution containing 5 mg/mL trypsin–EDTA (25300054, Sigma-Aldrich) at 37 °C. The dissociated cells were gently triturated to obtain a single-cell suspension. Neurons were plated at appropriate densities onto poly-D-lysine–coated culture plates in Minimum Essential Medium (11095080, Gibco) supplemented with penicillin–streptomycin, 10% FBS (SH30396.03HI, HyClone), 20% glucose and 200 mM L-glutamine (25030081, Gibco). After 1 hour of attachment, the plating medium was replaced with serum-free Neurobasal medium (21103049, Gibco) containing penicillin–streptomycin, 200 mM L-glutamine, and 2% B-27 supplement (17504044, Gibco) and. On DIV3, 4 µM cytosine β-D-arabinofuranoside (C1768, Sigma-Aldrich) was added to inhibit glial proliferation. Neurons were maintained in a humidified incubator at 37 °C with 5% CO₂, and immunoblotting experiments were performed on DIV14.

### Immunoblot analysis

Cells were lysed in ice-cold lysis buffer composed of 50 mM Tris–HCl (pH 7.5), 150 mM NaCl, 1 mM EDTA (46-034-Cl, Corning), 1.0% Triton X-100, and Halt™ EDTA-free Protease and Phosphatase Inhibitor Cocktail (78440, Thermo Fisher Scientific) for 10 minutes on ice. Lysates were centrifuged at 15,000 × g for 15 minutes at 4 °C, and the resulting supernatants were collected and kept on ice. Protein concentrations were determined using the Pierce™ BCA Protein Assay Kit (23227, Thermo Fisher Scientific) according to the manufacturer’s instructions. To analyze secreted proteins, culture medium was collected and centrifuged at 4,000 × g for 5 minutes to remove debris. The resulting supernatant was subjected to trichloroacetic acid (TCA) precipitation to concentrate proteins. Precipitated proteins were resuspended in SDS sample buffer at volumes normalized to the corresponding cell lysate protein concentrations, using the same buffer composition as for cell lysates. For protease protection assays, medium was first cleared by centrifugation as above and then incubated on ice for 2 hours in the presence or absence of 100 µg/mL proteinase K (25530049, Invitrogen) and 0.5% Triton X-100. Protease activity was subsequently quenched by addition of protease inhibitor cocktail, and proteins were recovered by TCA precipitation. Equal amounts of total protein were resolved by SDS–PAGE on 4–12% Bis-Tris gels (Invitrogen) and transferred onto 0.45 μm PVDF membranes (Millipore). Membranes were blocked with LI-COR Blocking Buffer (927-60001, LI-COR Biosciences) at room temperature and incubated with primary antibodies as indicated in the figure legends. For fluorescent detection, membranes were incubated with IRDye-conjugated secondary antibodies and imaged using a LI-COR Odyssey imaging system. For selected antibodies, HRP-conjugated secondary antibodies were used, and chemiluminescent signals were visualized with SuperSignal West Pico Plus Chemiluminescent HRP Substrate (34580, Thermo Scientific) and captured using a ChemiDoc Imaging System (Bio-Rad). Signal intensities from both detection methods were quantified using FIJI (ImageJ, NIH).

### Mouse urine collection

Urine was collected in clean cages without bedding, with 50 mL pipette basins placed evenly on the cage floor. Mice were restrained on the metal cage lid insert by placing the index finger and thumb on either side of the neck and the palm over the body. Most mice urinated spontaneously, without requiring additional massage to the abdomen or back. If no urine was produced, the collection was repeated on another day. Approximately 50-100 µL of urine was collected at a time. Samples were collected by pipette and stored at -80°C until analysis. All collections were performed at a consistent time of day in mice aged 6-8 months.

### Tissue homogenization

Brain and kidney samples were processed using a mechanical dounce homogenizer. Tissues were kept on ice in sucrose homogenization buffer (0.32M sucrose, 1mM NaHCO3, 20 mM HEPES, 1mM MgCl2, 1% Halt™ Protease and Phosphatase Inhibitor Cocktail (ThermoFisher, #78440)) and plunged 20 times. Lysed tissue was spun down at 1000g for 10 mins at 4°C. The resulting supernatant was transferred and spun again at 1000g for 10 mins. The final supernatant was stored at -80°C until analysis.

### qRT-PCR

Complimentary DNA (cDNA) synthesis and quantitative real-time PCR (qPCR) were performed as previously described (Choi I, et al. Nat Cell Biol. 2023 Jul;25(7):963-974. doi: 10.1038/s41556-023-01158-0. Epub 2023 May 25. PMID: 37231161; PMCID: PMC10950302. --add to citation) Briefly, cDNA was synthesized using the AffinityScript Multi-Temp Reverse Transcriptase (Stratagene) and oligo (dT)_18_ primer. qPCR was performed using PlatinumTaq DNA polymerase (Invitrogen) with SYBR green (Molecular Probes) detection, using an ABI7900HT thermocycler (Applied Biosystems). The PCR conditions used were: 95 °C for 2 min, 40 cycles of 95 °C for 15 s, 55 °C for 15 s, and 72 °C for 30 s for the following primer pairs (Eurofins MWG Operon):

Human LRRK2: CAGATTGCCCCTGACTTGAT (sense), TCATAGGCTGCTCGGTAAAC (antisense)

Mouse LRRK2: TAAATCCAGACCAACCAAGGCTC (sense), TCC TTC GTA GGC AGC TCG AT (antisense);

Actin: 5′-AGGTGACAGCATTGCTTCTC-3′ (sense), 5′-GCTGCC TCAACACCTCAAC-3′ (antisense)

Relative gene expressions were quantified using the 2^-ΔΔCt^ method.

### Human CSF and urine proteomics for DAPs analysis and the human CSF longitudinal proteomics

The human CSF proteomic data were measured using the Slow Off-rate Modified Aptamer (SOMA)scan platform. For data quality control, the outlier samples, calibrators, buffer and non-human SOMAmers were removed. The raw measured values were hybridization normalized, plate scaled, median normalized intra plate and calibrated at SomaLogic’s side, following log2 transformed, median normalized inter plates and batch corrected at plate level. More details about the data process protocol are available elsewhere (https://www.ppmi-info.org/access-data-specimens/download-data).

For the human urine, proteins were extracted from urine samples and subsequently digested into peptides through the utilization of the MStern blotting sample preparation protocol^12,49^. The protocol underwent a minor modification and was semi-automated on an Agilent Bravo liquid handling system. For urinary proteome profiling, the purified peptide mixtures were applied to Evosep Evotips and subjected to nanoflow separation on an Evosep One HPLC^50^. Chromatographic separation was carried out over a 44-minute gradient, and the eluted peptides were analyzed on a Bruker timsTOF Pro mass spectrometer using a data-independent acquisition (DIA) method^51^. More details about the data process protocol are available elsewhere (https://www.ppmi-info.org/access-data-specimens/download-data).

Human longitudinal proteomic profiling was performed on cerebrospinal fluid (CSF) from participants enrolled in the Parkinson’s Progression Marker Initiative (PPMI) cohort, including individuals with Parkinson’s disease and healthy controls. A total of 2,283 CSF samples derived from 482 participants were analyzed by data-independent acquisition mass spectrometry. Raw MS data were processed using the OpenSWATH pipeline, followed by quantitative processing with mapDIA and subsequent batch-correction procedures. More details about the data process protocol are available elsewhere (https://www.ppmi-info.org/access-data-specimens/download-data).

### Mouse urine sample preparation

Mouse urine samples were processed using a modified urine-HILIC protocol^52,53^. Briefly, 40 µL of urine was transferred into 1.5 mL Eppendorf tubes containing three volumes (120 µL) of lysis buffer (8 M urea, 2% SDS in 50 mM Tris-HCl, pH 8.0). Samples were reduced by adding 4 µL of 410 mM dithiothreitol (DTT in 50 mM Tris-HCl, pH 8.0) to obtain a final concentration of 10 mM and incubated for 30 min at room temperature. Alkylation was performed by adding 5.2 µL of 960 mM chloroacetamide (CAA in 50 mM Tris-HCl, pH 8.0) to obtain a final concentration of 30 mM, followed by incubation for another 30 min in the dark at room temperature. Separately, MagReSyn® HILIC beads (ReSyn Biosciences) were prepared by washing twice with equilibration buffer (15% acetonitrile [ACN] in 100 mM ammonium acetate, pH 4.5) and resuspending in the same buffer to achieve a final bead suspension of 20 µg/µL. After lysis, reduction, and alkylation, each urine lysate was mixed 1:1 (v/v) with binding buffer (30% ACN in 200 mM ammonium acetate, pH 4.5), and 4 µL of equilibrated bead suspension was added (bead-to-sample ratio 1:10, v/v). The mixture was gently agitated at 700 rpm for 30 min at room temperature to promote protein binding. Beads were collected magnetically, washed twice with 95% ACN, and resuspended in 100 µL digestion buffer (100 mM triethylammonium bicarbonate [TEAB], pH 8.5). Bound proteins were digested overnight at 37 °C with trypsin and LysC (0.2 µg each; 1:50 enzyme-to-protein ratio, w/w). Peptide concentrations were determined at 280 nm using a NanoDrop 2000 spectrophotometer (Thermo Scientific). For LC-MS analysis, 500 ng of peptide material was loaded onto Evotips (Evosep Biosystems) following the manufacturer’s protocol. Briefly, Evotips were activated with 1-propanol, washed with 0.1% formic acid (FA) in ACN, equilibrated with 0.1% FA, loaded with samples, washed again with 0.1% FA, and stored in 150 µL of 0.1% FA to prevent drying prior to timsTOF Pro 2 analysis.

### Data acquisition

LC-MS/MS analysis was performed on an Evosep One liquid chromatography system (Evosep Biosystems) coupled to a timsTOF Pro 2 mass spectrometer (Bruker Daltonics). Purified peptides were analyzed using the 30 samples per day (30SPD) method, featuring a 44-minute active gradient with a mobile phase composed of 0.1% formic acid in LC/MS-grade water (buffer A) and 0.1% formic acid in acetonitrile (buffer B). Peptides were separated on a PepSep analytical column (15 cm × 75 µm ID, 1.5 µm C18; Bruker Daltonics) maintained at 50 °C and connected to a fused-silica emitter (10 µm ID) operated within a nanoelectrospray ion source (CaptiveSpray, Bruker Daltonics). The timsTOF Pro 2 was operated in dia-PASEF mode using a method optimized with the Python tool py_diAID to maximize precursor coverage of tryptic digests^54^. The final method comprised one MS1 scan followed by twenty dia-PASEF scans, each containing two ion-mobility (IM) ramps, covering an m/z range of 350-1,200 and an IM range of 0.7-1.3 Vs cm⁻². The collision energy increased linearly from 20 eV at 1/K₀ = 0.6 Vs cm⁻² to 59 eV at 1/K₀ = 1.6 Vs cm⁻².

### Raw data analysis with DIA-NN

DIA-NN version 1.9^55^ was used to perform a library-free search on the dia-PASEF files against the Mus musculus UniProt FASTA database (taxonomy ID 10090; 17,222 protein entries and 16,878 gene entries, downloaded in January 2025). The spectral library was generated through an in silico tryptic digestion, allowing a maximum of one missed cleavage. Cysteine carbamidomethylation was set as a fixed modification, and methionine oxidation was selected as a variable modification, with the maximum number of variable modifications restricted to one. The MS1 mass accuracy was configured to 10 ppm, and the MS2 mass accuracy was adjusted to 15 ppm. The scan window radius was set to 7. Match-between-runs was also enabled within the search. All other settings were kept at default.

For the human urine and mouse urine proteomic datasets, we used Perseus software package versions 2.1.3.0^56^ to do the log2 transformation, low abundance protein filtering (>70%) and imputation with normal distribution.

### Differentially abundant protein analysis

For all these datasets, the significance of differential abundance was evaluated based upon statistical criteria through R package limma^57^ (version 3.64.3), with FDR < 0.05 and Log2(FC) > 1.5SD, or *P*-value < 0.05 and Log2(FC) > 1.5SD. For human CSF and urine proteomic datasets, we also adjusted with the age and sex and the outliers were identified and excluded through Principle Component Analysis (PCA). The SD of proteins was estimated by fitting to a Gaussian distribution to evaluate the magnitude of experimental variations.

### Machine learning

OmicLearn (v1.4)^58^ was utilized for performing data analysis, model execution, and creation of plots and charts. Log2 transferred and normalized protein expression intensity tables were used as the input for OmicLearn. Machine learning was done in Python (3.10.11). Feature tables were imported via the Pandas package (1.5.3) and manipulated using the Numpy package (1.24.2). The machine learning pipeline was employed using the scikit-learn package (1.2.2). The Plotly (5.9.0) library was used for plotting. No normalization on the data was performed. To impute missing values, a Zero-imputation strategy was used. Features were selected using a ExtraTrees (n_trees=100) strategy with a maximum number of 30 features. 70% of the samples were randomly selected for training and the rest 30% were used for testing, during the training, normalization and feature selection was individually performed using the data of each split. For classification, RandomForest-Classifier (random_state = 23, n_estimators = 100, criterion = gini, max_features = auto) was used. When using a repeated (n_repeats=10), stratified cross-validation (RepeatedStratifiedKFold, n_splits=5) approach to classify LRRK2 PD vs. HC and sPD vs. HC in human CSF, as well as LRRK2 PD vs. HC in human urine. The average feature importance scores assigned by the classifier for each of the top20 features are shown in **Fig. 1d**, and **Fig. 4c**.

### Pathway enrichment analysis

GO term and KEGG were performed with the R Bioconductor package clusterProfiler^59^. Briefly, we first transfer the proteins symbols to Entrez IDs based on Bioconductor annotation data package (org.Hs.eg.db/org.Mm.eg.db) and used the clusterProfiler::enrichGO with the pvalueCutoff = 0.05, qvalueCutoff = 0.05 to do pathway enrichment analysis. For KEGG analysis, we used clusterProfiler::enrichKEGG with the pvalueCutoff = 0.05. All the volcano plots were generated by EnhancedVolcano^60^ R package.

### Weighted gene correlation network analysis (WGCNA)

The analysis was carried out with the WGCNA^61^ R package. For the human CSF proteomic data, we used the blockwiseModules function to generate the network, with the settings soft-threshold power = 10, networkType = “signed”, minModuleSize = 30, deepSplit = 3, reassignThreshold = 1e-3, mergeCutHeight = 0.2, numericLabels = F, pamRespectsDendro = FALSE, saveTOMs = TRUE, randomSeed = 1234, maxBlockSize = 20000, and verbose = 3. The partitioning around medoids-like (PAM-like) stages of module detection was disabled to ensure that module assignments adhered strictly to the hierarchical clustering dendrogram (pamRespectsDendro = FALSE). The soft-threshold power of 10 was chosen based on the fitted signed scale-free topology model (R2) approaching an asymptote of 0.85 at that threshold. Furthermore, the mean connectivity at that power was less than 100. The maximum block size was set at 20,000 to complete all clustering within a single block. For the human urine proteomic data, the settings were: soft-threshold power = 12, networkType = “signed”, minModuleSize = 30, deepSplit = 3, reassignThreshold = 1e-3, mergeCutHeight = 0.2, numericLabels = F, pamRespectsDendro = FALSE, saveTOMs = TRUE, randomSeed = 1234, maxBlockSize = 20000, and verbose = 3. For the mouse urine proteomic data, the settings were: soft-threshold power = 14, networkType = “signed”, minModuleSize = 30, deepSplit = 3, reassignThreshold = 1e-3, mergeCutHeight = 0.2, numericLabels = F, pamRespectsDendro = FALSE, saveTOMs = TRUE, randomSeed = 1234, maxBlockSize = 20000, and verbose = 3.

### Protein-protein interaction (PPI) network analysis

The protein-protein interaction network analysis was performed using the STRING^30^ plugin in CytoScape^62^, with the fold changes and FDR/*P*-values obtained from the differentially expressed proteins analysis. Briefly, for the human and mouse proteomic datasets in M4, M10, and M9, we analyzed the PPI network among these proteins using STRING. Subsequently, we adjusted the node colors and sizes based on the fold changes and FDR/*p*-value obtained from differentially abundant protein analysis (human CSF LRRK2 PD vs. HC, human urine LRRK2 PD vs. HC, and mouse urine h*LRRK2*^G2019S^ vs. WT).

### Linear mixed effect model

To analyze the longitudinal data collected during follow-up, we applied linear mixed models to investigate the linear associations between variables:

Y = X β + Z μ + ε

where Y is a vector of observations, β represents fixed effects, μ represents random effects and ε is a vector of random errors. X and Z are model matrices of independent variables related to β and μ, respectively.

In this study, we applied random intercepts for participant ID to account for within-subject variations across repeated measurements. To identify protein trajectories that exhibited significant longitudinal trends, we used the following model specification:

Protein level ∼ α + β1 group + β2 time + β3 group: time + β4 age + (1 | participant ID)

Thus, protein levels were modeled as a function of follow-up time (since symptom onset), group, their interaction, and age (centered), with participant-specific random intercepts to account for individual baseline variation. The follow-up time was defined relative to symptom onset rather than diagnosis.

Individuals lacking a reported onset time were excluded from the analysis. In addition, one LRRK2 PD sample (PATNO: 40585) was removed from further analysis because it exhibited multiple protein intercepts that differed from the cohort mean by over two standard deviations.

### Ingenuity pathway analysis (IPA)

The canonical and upstream regulator analysis within comparison analysis were performed through QIGEN IPA software (version 01-23-01)^36^. We first did the differentially abundant protein analysis of different comparison, then we analyzed these results individually with the core analysis modular in IPA. Lastly, we input all the results generated by core analysis and did the comparison analysis. We then downloaded the top 10 canonical and upstream regulator results and plotted the results with a R package “heatmap” (ref), shown as **Supplementary Fig.** 9b,c. The radial plots were generated directly from IPA.

### Statistical analysis

The statistical significance of the difference between the groups was determined using a One-Way ANOVA followed by Tukey’s multiple comparisons test unless otherwise specified. The latest version of GraphPad Prism 10 (GraphPad Software) was used for statistics and graph images. No statistical method was used to pre-determine the sample size. No data were excluded from the analyses. All values are reported as mean ± standard error of the mean (s.e.m.)

### List of Supplementary Materials

Figs. S1 to S9

Tables S1 to S31

## Data Availability

All data produced in the present study are available upon reasonable request to the authors
All data produced in the present work are contained in the manuscript
All data produced are available online at https://www.ppmi-info.org/access-data-specimens/download-data

## Acknowledgements

The schematics were created using BioRender.

## Author contributions

Conceptualization: ZY, CX, TS

Methodology: CX, TS, BTH

Investigation: CX, TS, BTH

Visualization: CX, TS, BTH

Funding acquisition: ZY

Project administration: ZY, CX, TS

Supervision: ZY, CX, TS

Writing – original draft: ZY, CX, TS, BTH

Writing – review & editing: ZY, CX, TS, BTH, OK, DTV

## Data availability

The authors affirm that all supporting data for the study’s findings are readily available within the paper and its Supplementary Information files. The mass spectrometry proteomics data have been deposited to the ProteomeXchange Consortium via the PRIDE^63^ partner repository with the dataset identifier . For further inquiries regarding resources and reagents, please contact the lead contact, Zhenyu Yue. The source data accompanying this publication are also provided.

---

**Supplementary Fig. 1.**
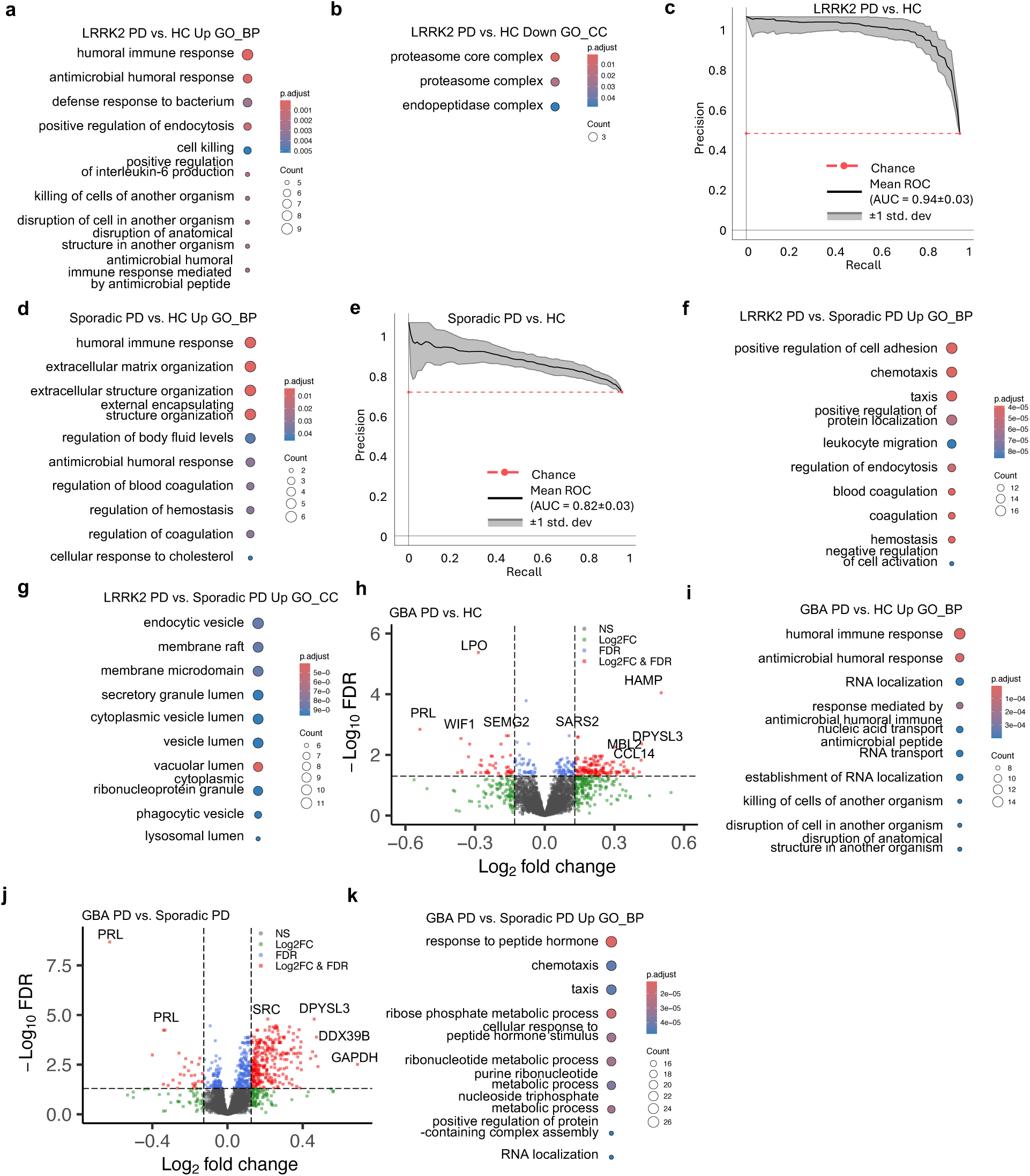
Analysis of CSF proteomics from GBA PD, sporadic PD, and healthy controls. **a,b,d,f,g** Gene Ontology (GO) annotations of the DAPs in **Fig. 1b** (**a,b**), **Fig. 1f** (**c**), and **Fig. 1j** (**d**,**e**), displaying the top 10 enriched pathways (*P*-value < 0.05, one-sided hypergeometric test). **c,e,** Precision–Recall (PR) curve and corresponding AUC statistics generated from 5-fold cross-validation repeated 10 times using the RandomForest-based model to classify LRRK2 PD (**c**) /sPD (**d**) vs. healthy controls based on the feature protein panel. The black line represents the mean PR curve across all repeats, and the gray band denotes the ±1 standard deviation interval. The red dashed line indicates the expected performance under random chance, corresponding to a classifier with no discriminative ability. Mean AUC and standard deviation are shown in parentheses. **h,j,** Volcano plot of the differentially abundant proteins (DAPs) detected by mass-spectrometry in human CSF for GBA PD vs. HC (**h**) and GBA PD vs. sPD (**j**). A positive score indicates enrichment, a negative score indicates depletion. The y axis represents statistical confidence FDR (adjusted *P*-value) for each x axis point. Enriched proteins, defined by FDR < 0.05 and |Log2FC| > 1.5SD are colored in red. FDR was calculated with the moderated two-sided *t*-test in the LIMMA package for multiple comparisons. **i,k,** Gene Ontology (GO) annotations of the DAPs in **Supplementary Fig. 1 h** (**i**), **Supplementary Fig. 1j** (**k**), displaying the top 10 enriched pathways (*P*-value < 0.05, one-sided hypergeometric test).

**Supplementary Fig. 2.**
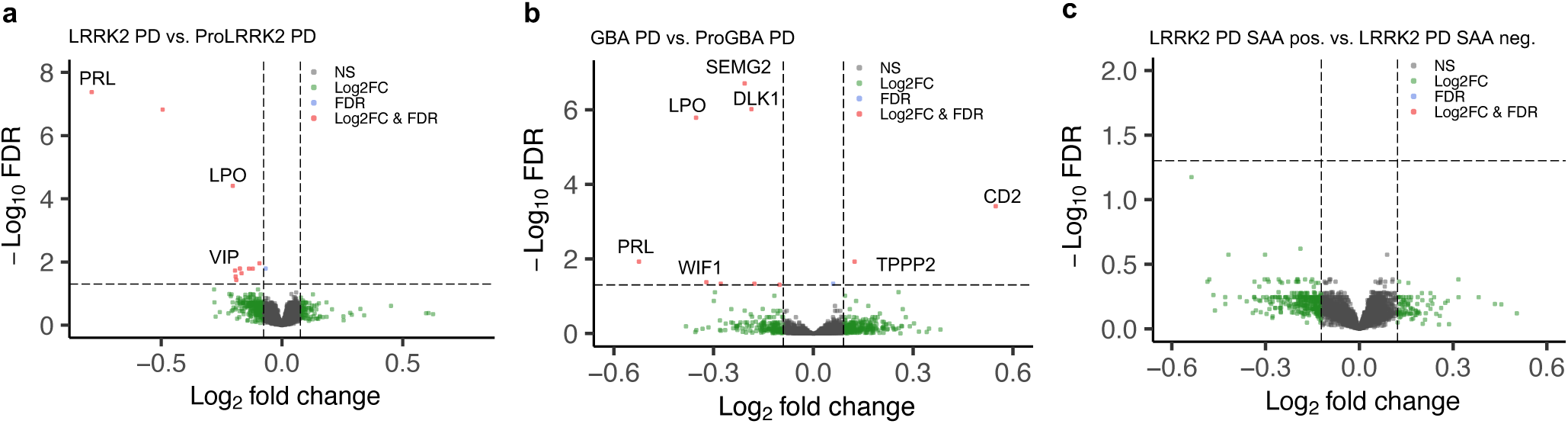
Analysis of CSF proteomics in disease phases and subgroups of LRRK2 and GBA PD populations. **a,b,c,** Volcano plot of the differentially abundant proteins (DAPs) detected by mass-spectrometry in human CSF for LRRK2 PD vs. prodromal LRRK2 PD (**a**) and GBA PD vs. prodromal GBA PD (**b**), and LRRK2 PD SAA positive vs. LRRK2 PD SAA negative. A positive score indicates enrichment, a negative score indicates depletion. The y axis represents statistical confidence FDR (adjusted *P*-value) for each x axis point. Enriched proteins, defined by FDR < 0.05 and |Log2FC| > 1.5SD are colored in red. FDR was calculated with the moderated two-sided *t*-test in the LIMMA package for multiple comparisons.

**Supplementary Fig. 3.**
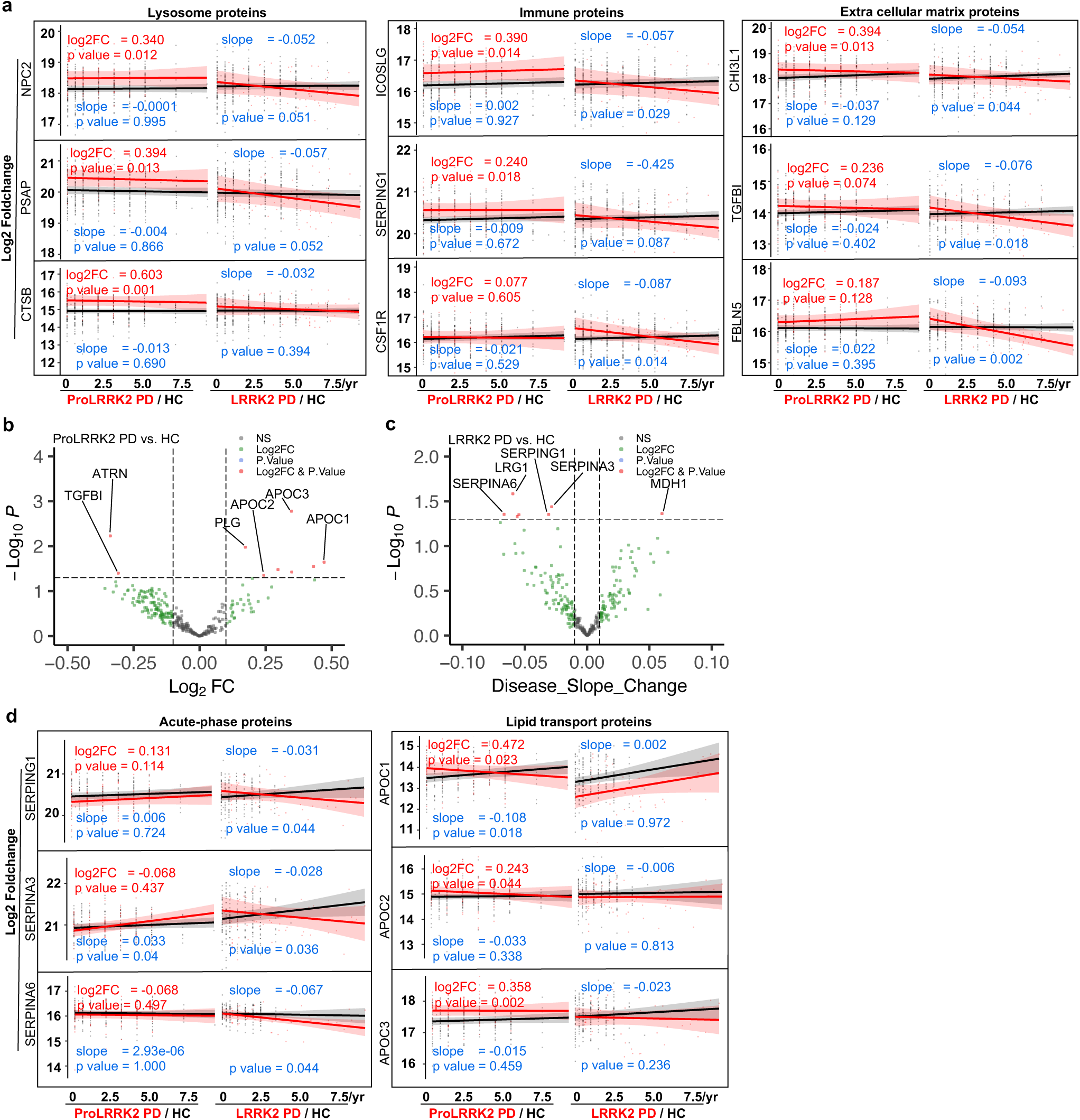
Analysis of longitudinal CSF datasets of LRRK2 PD and healthy control. **a**, Dot plots of longitudinal trajectories of selected protein abundances across disease progression in males. Each dot represents the specific protein expression in an individual sample. The red line denotes the estimated fixed-effect trajectory for prodromal LRRK2 PD or LRRK2 PD, and the black line represents HC. Shaded ribbons indicate the 95% confidence intervals (CIs) of the fixed-effect mean trajectories. Corresponding log2FC, slopes, and *P*-values are shown alongside each trajectory. **b**, Volcano plot of the significant enriched proteins at baseline detected by mass-spectrometry in human CSF for prodromal LRRK2 PD vs. HC in females. A positive score indicates enrichment, a negative score indicates depletion. The y axis represents statistical confidence *P*-value for each x axis point. Enriched proteins, defined by *P*-value < 0.05 and |Log2FC| > 0.1 are colored in red. *P*-value was calculated with the moderated two-sided *t*-test in the LIMMA package. **c**, Volcano plot of the proteins with significant slopes detected by mass-spectrometry in human CSF for LRRK2 PD vs. HC in females. A positive score indicates an increase, a negative score indicates a decrease. The y axis represents statistical confidence *P*-value for each x axis point. Proteins with significant slope change, defined by *P*-value < 0.05 and |slope| > 0.01 are colored in red. *P*-value was calculated with the moderated two-sided *t*-test in the LIMMA package. **c**, Dot plots of longitudinal trajectories of selected protein abundances across disease progression in females. Each dot represents the specific protein expression in an individual sample. The red line denotes the estimated fixed-effect trajectory for prodromal LRRK2 PD or LRRK2 PD, and the black line represents HC. Shaded ribbons indicate the 95% confidence intervals (CIs) of the fixed-effect mean trajectories. Corresponding log2FC, slopes, and *P*-values are shown alongside each trajectory.

**Supplementary Fig. 4.**
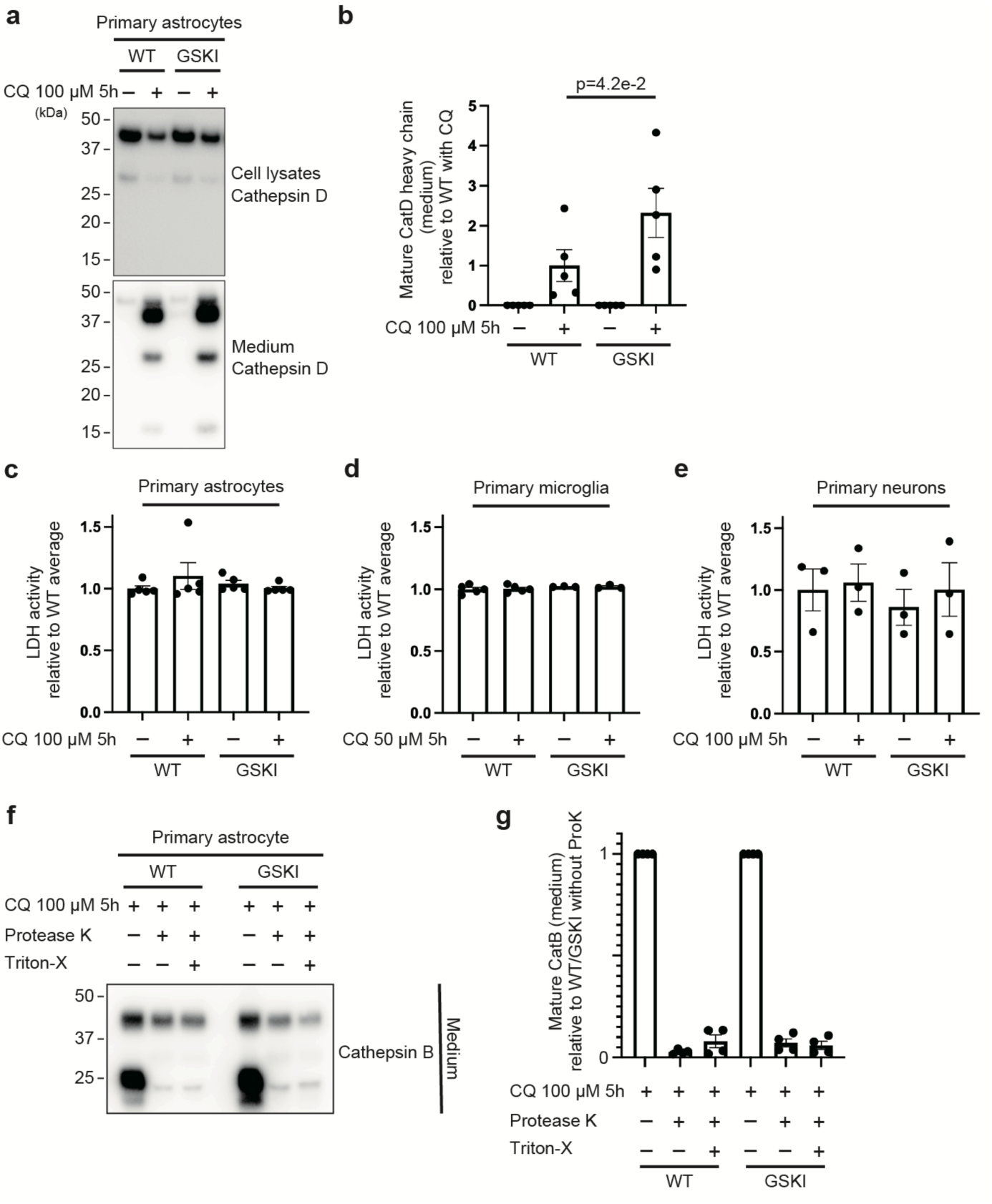
Extended analyses of lysosomal secretion in *Lrrk2* mutant glia and neurons. **a,** Cell lysates and conditioned media from primary astrocytes treated with or without 100 μM chloroquine (CQ) for 5 hours were subjected to immunoblot analysis using the indicated antibodies. Primary astrocytes were prepared from five independent wild-type (WT) or *Lrrk2*^G2019S^ knock-in (GSKI) pups. **b,** Quantification of mature Cathepsin D heavy chain levels in the conditioned media shown in (**a**). Values were normalized to the average of WT + CQ condition. Each data point represents an independent biological replicate derived from a distinct pup (astrocytes: n = 5; neurons: n = 3). Bars indicate the mean ± standard error of the mean (SEM). Statistical significance was determined by one-way ANOVA followed by Holm–Sidak post hoc test. **c,d,e,** LDH activity in the culture media of primary astrocytes (**c**), primary microglia (**d**) and primary neurons (**e**) derived from WT or GSKI pups, treated with or without chloroquine (CQ) at the indicated concentrations for 5 hours. LDH activity is shown relative to the average of the WT control group. Data represent mean ± SEM (n = 5 for astrocytes, n = 5 for WT microglia and n =3 for GSKI microglia, n = 3 for neurons). **f,** Conditioned media from primary astrocytes derived from WT or GSKI pups treated with 100 μM chloroquine (CQ) for 5 hours were subjected to proteinase K (ProK) treatment (with or without 0.5% Triton X-100) for 3 hours, followed by immunoblotting for Cathepsin B. **g,** Quantification of mature Cathepsin B levels in the media under each condition, normalized to the no-ProK control for each genotype. Each dot represents an independent biological replicate derived from a distinct pup (n = 4).

**Supplementary Fig. 5.**
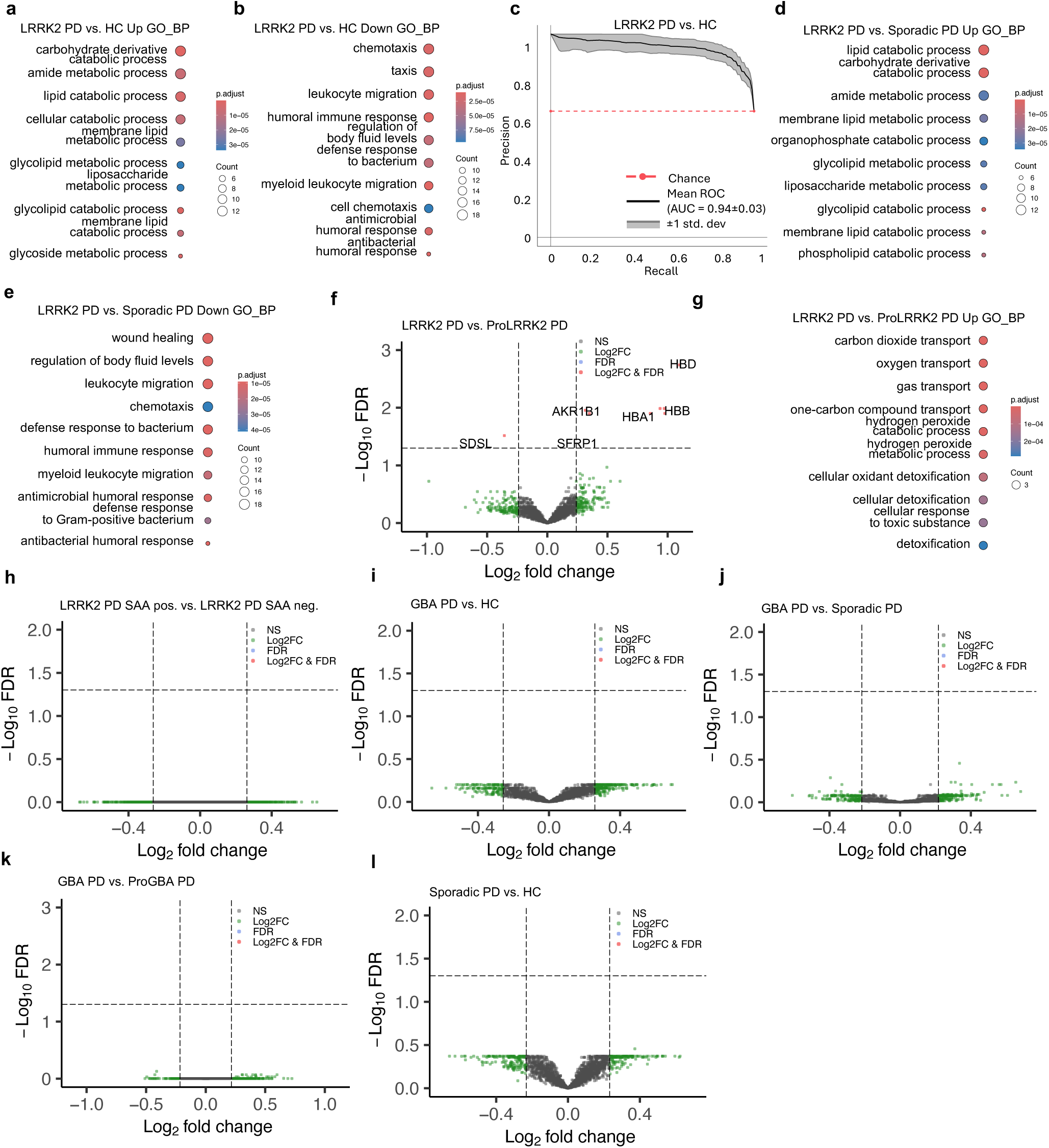
Analysis of urine proteomics from GBA PD, sporadic PD, and healthy controls. **a,b,d,e,** Gene Ontology (GO) annotations of the DAPs in **Fig.** 4**a** **(a,b)** and **Fig.** 4**e** **(d,e)**, displaying the top 10 enriched pathways (*P*-value < 0.05, one-sided hypergeometric test). **c,** Precision–Recall (PR) curve and corresponding AUC statistics generated from 5-fold cross-validation repeated 10 times using the RandomForest-based model to classify LRRK2 PD vs. healthy controls based on the feature protein panel. The black line represents the mean PR curve across all repeats, and the gray band denotes the ±1 standard deviation interval. The red dashed line indicates the expected performance under random chance, corresponding to a classifier with no discriminative ability. Mean AUC and standard deviation are shown in parentheses. **f,h,i,j,k,l,** Volcano plot of the differentially abundant proteins (DAPs) detected by mass-spectrometry in human urine for LRRK2 PD vs. prodromal LRRK2 PD (**e**), LRRK2 PD SAA positive vs. LRRK2 PD SAA negative (**g**), GBA PD vs. HC (**h**), GBA PD vs. sPD (**i**), and GBA PD vs. prodromal GBA PD (**j**). A positive score indicates enrichment, a negative score indicates depletion. The y axis represents statistical confidence FDR (adjusted *P*-value) for each x axis point. Enriched proteins, defined by FDR < 0.05 and |Log2FC| > 1.5SD are colored in red. FDR was calculated with the moderated two-sided *t*-test in the LIMMA package for multiple comparisons. **g,** Gene Ontology (GO) annotations of the DAPs in **Supplementary Fig. 1 f** (**g**), displaying the top 10 enriched pathways (*P*-value < 0.05, one-sided hypergeometric test).

**Supplementary Fig. 6.**
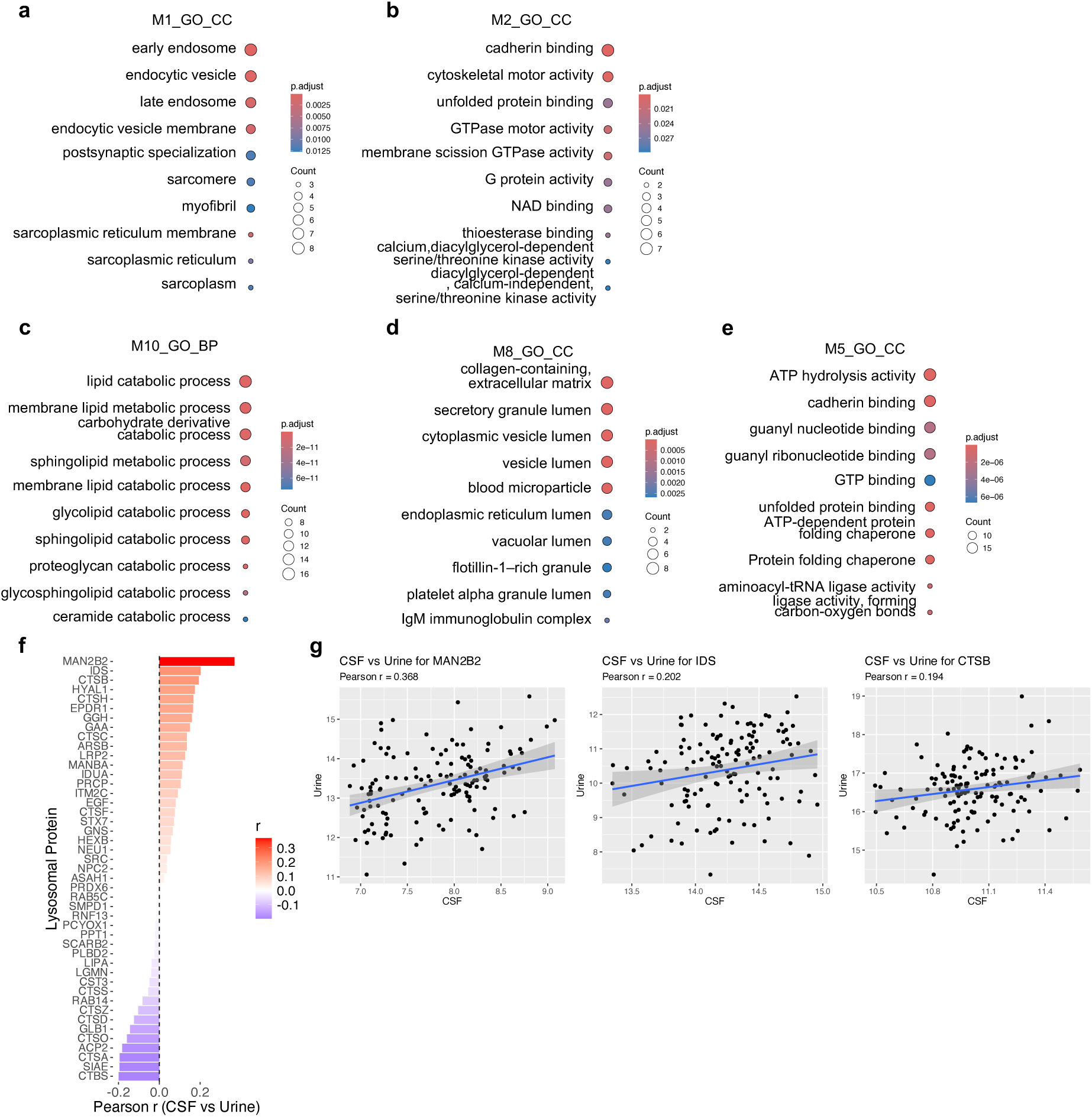
Analysis of urine proteomics from GBA PD, sporadic PD, and healthy controls. **a,b,c,d,e,** Gene Ontology (GO) annotations of the proteins in M1 (**a**) and M2 (**b**) from human CSF and in M10 (**c**), M8 (**d**), and M5 (**e**) from huma urine, displaying the top 10 enriched pathways (*P*-value < 0.05, one-sided hypergeometric test). **f,** The bar plot of *Pearson* correlations of the detected lysosomal core proteins in both CSF and urine from the same LRRK2 PD individuals. The color gradience represents from low (blue) to high (red) scores. **g,** The scatter plot of top 3 proteins in **Supplementary Fig. 6f**. The x axis represents the log2 transferred human CSF protein intensity values, the y axis represents the log2 transferred human urine protein intensity values.

**Supplementary Fig. 7.**
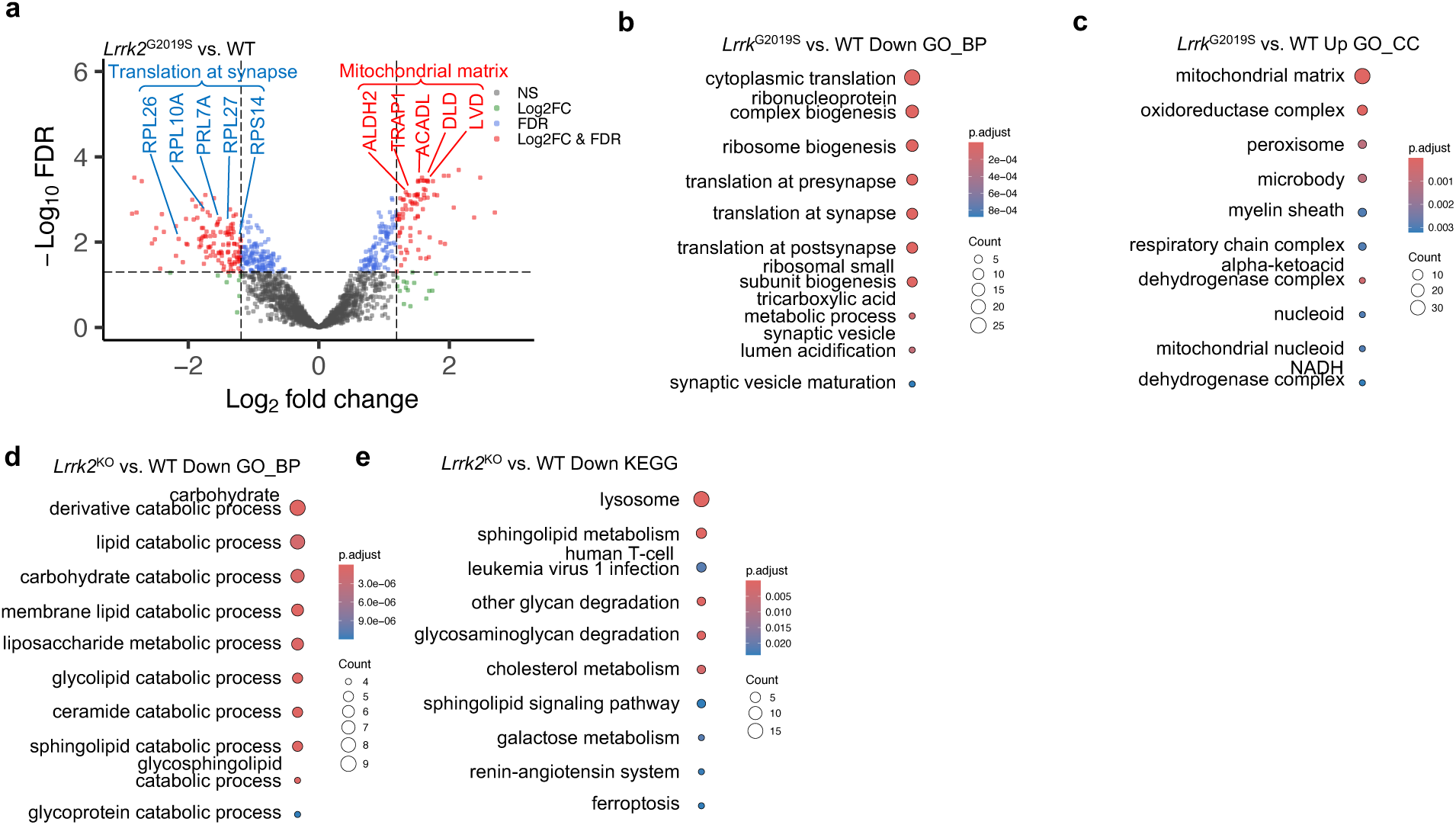
The enriched pathways of the comparative urine DEP analysis. **a,** Volcano plot of the differentially abundant proteins (DAPs) detected by mass-spectrometry in mouse urine for *Lrrk2*^G2019S^ vs. WT. A positive score indicates enrichment, a negative score indicates depletion. The y axis represents statistical confidence FDR (adjusted *P*-value)/*P*-value for each x axis point. Enriched proteins, defined by FDR/*P*-value < 0.05 and |Log2FC| > 1.5SD are colored in red. FDR/*P*-value was calculated with the moderated two-sided *t*-test in the LIMMA package for multiple comparisons. **b,c,d,** Gene Ontology (GO) annotations of the DAPs in Supplementary **Fig. 7a** (**b,c**) and **Fig. 6a** (**d**), displaying the top 10 enriched pathways (*P*-value < 0.05, one-sided hypergeometric test). **e,** Kyoto Encyclopedia of Genes and Genomes (KEGG) annotations of the DAPs in **Fig. 6a** (**e**), displaying the top 10 enriched pathways (*P*-value < 0.05, one-sided hypergeometric test).

**Supplementary Fig. 8.**
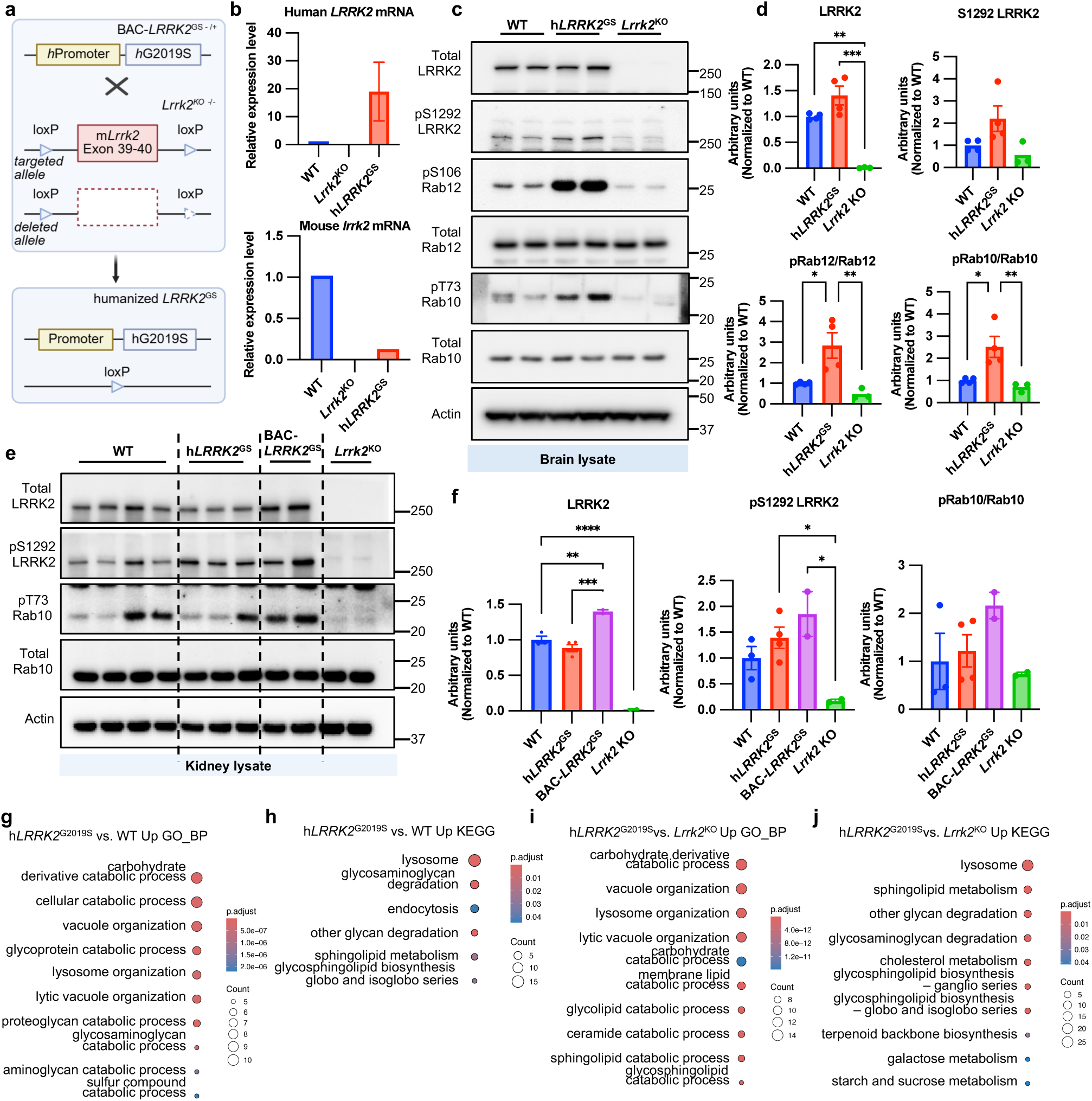
Validation of human LRRK2 expression and function in humanized hLRRK2 GS model. **a,** Breeding schematic showing BAC-h*LRRK2*^G2019S^ mice containing the full human LRRK2 sequence with its promoter was crossed with *Lrrk2*^KO^ to generate the humanized LRRK2^G2019S^ mice line. **b,** Comparison of human *LRRK2* and mouse *lrrk2* mRNA expression in brain tissues from h*LRRK2*^G2019S^, *Lrrk2*^KO^ and WT mice using RT-qPCR. **c,e,** Immunoblot analysis of whole brain (n= 3-4) (**c**) and kidney lysates (n= 2-4) (e) from 6 months old male mice across genotypes. **d,f,** Quantification of brain d) and kidney f) immunoblots showing total LRRK2 and the phosphorylation of its downstream substrates, Rab12 and Rab10, normalized to actin. Statistical analysis was performed by one-way ANOVA with post hoc Tukey’s test (****P <0.0001, ***P <0.001, **P<0.01, *P<0.05; SEM bars shown) **g,i,** Gene Ontology (GO) annotations of the DAPs in **Fig. 6c** (**g**), **Fig. 6e** (**i**), displaying the top 10 enriched pathways (*P*-value < 0.05, one-sided hypergeometric test). **h,j,** Kyoto Encyclopedia of Genes and Genomes (KEGG) annotations of the DAPs in **Fig. 6c** (**h**), and **Fig. 6e** (**j**), displaying the top 10 enriched pathways (*P*-value < 0.05, one-sided hypergeometric test).

**Supplementary Fig. 9.**
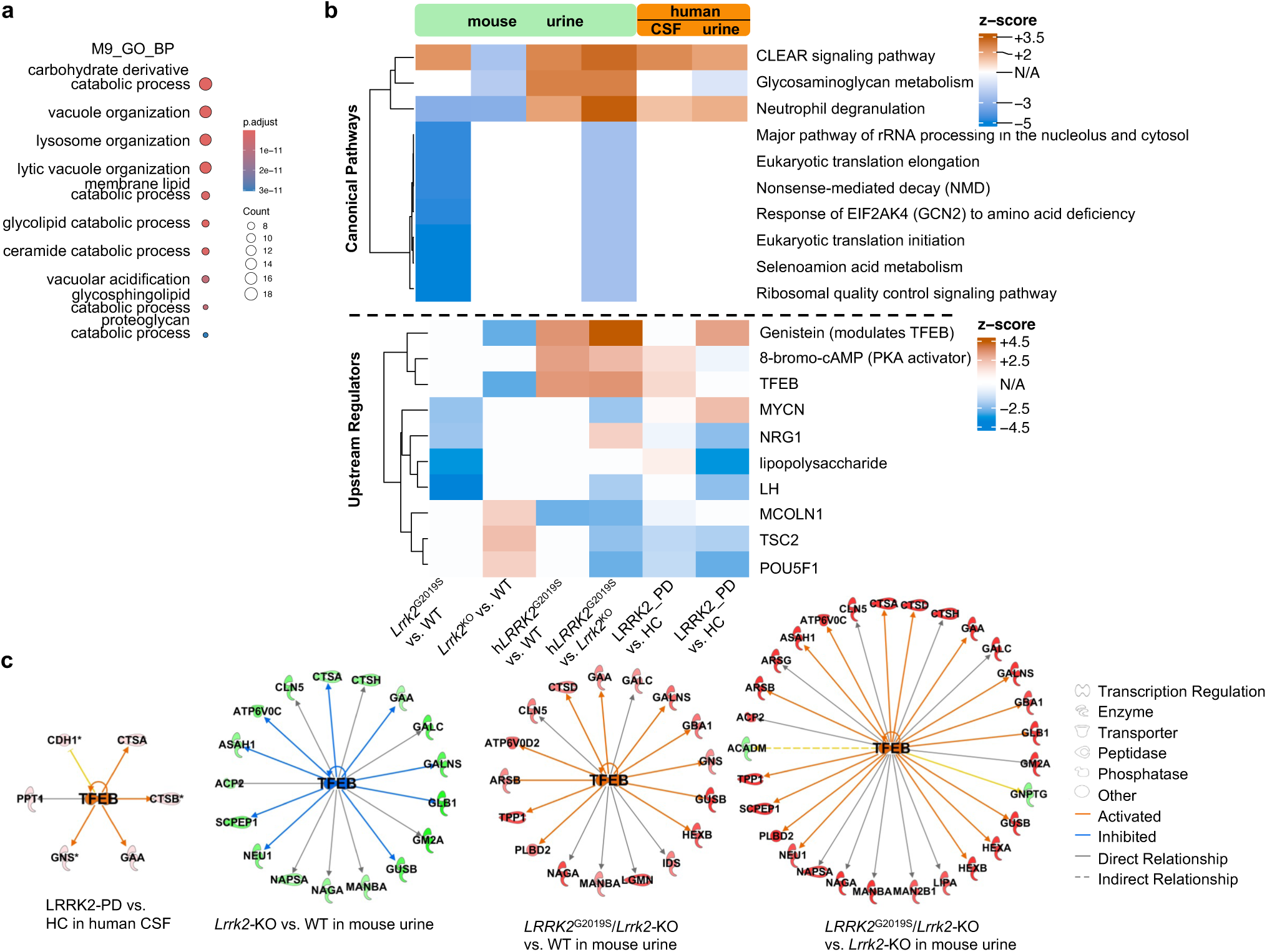
Integrated analysis of network modules and pathways across human and mouse biofluid datasets. **a,** Gene Ontology (GO) annotations of the proteins in M9 from mouse urine, displaying the top 10 enriched pathways (*P*-value < 0.05, one-sided hypergeometric test). **b,** Canonical pathway and upstream regulator analyses based on Ingenuity Pathway Analysis (IPA) across mouse urine, human CSF, and human urine datasets. Heatmaps depict the predicted activation (brown) or inhibition (blue) of pathways and regulators, with color intensity corresponding to activation z-scores. For canonical pathways (top), a z-score > 2 indicates predicted activation, and a z-score < −2 indicates predicted inhibition. For upstream regulators (bottom), hierarchical clustering was applied to highlight shared and distinct regulatory patterns across species and biofluids. **c,** Predicted TFEB regulatory networks in human CSF and mouse urine derived from Ingenuity Pathway Analysis (IPA). Network diagrams illustrate TFEB-associated lysosomal and metabolic targets identified in different LRRK2-related comparisons. In human CSF (top left), TFEB is predicted to be activated in LRRK2-PD relative to healthy controls (HC). In mouse urine (top right and bottom panels), TFEB-regulated networks are shown for *Lrrk2*^KO^ vs. WT, *LRRK2*^G2019S^ vs. WT, and *LRRK2*^G2019S^ vs. *Lrrk2*^KO^. Orange and blue lines indicate predicted activation and inhibition, respectively; solid lines denote direct and dashed lines indirect relationships. Node shapes represent molecular functions (e.g., enzyme, transporter, phosphatase), and node colors indicate activation status based on z-score predictions.

